# Global Health Injustice From Climate Change Driven By Consumption

**DOI:** 10.64898/2026.06.10.26355381

**Authors:** Lea Rupcic, Daniel Yoo, Annie Levasseur, Charles Alexandre, Alexis Laurent, Olivier Jolliet

**Author notes:** Equal contribution (Lea Rupcic, Daniel Yoo).

## Abstract

Climate change imposes unequal health burdens from heat and cold, disproportionately harming vulnerable nations least responsible for emissions. A framework to quantitatively attribute this damage to different countries’ consumption patterns has been missing. We developed a global framework linking consumption-based greenhouse gas emissions to country-specific health burdens, measured in Disability-Adjusted Life Years (DALYs). Our results quantify the profound scale of this externalized harm. For example, average North American consumption imposes a global health burden of 34 days of healthy life per person per year, without net damage suffered. In contrast, Sub-Saharan Africa endures 25 days per person per year despite minimal emissions. The resulting Health Injustice Index provides a powerful instrument for climate accountability, reframing responsibility in terms of tangible human health impacts.

## Main Text

Human-induced climate change imposes a severe and deeply unjust toll on global health (*1–4*). Rising global temperatures drive increases in mortality and morbidity, often characterized by more frequent and intense heatwaves and subsequent impacts on human health (*2*, *5*). The resulting human health burden—quantified comprehensively and comparably across diverse outcomes using Disability-Adjusted Life Years (DALYs) (*6*), a commonly used metric for capturing mortality and morbidity, is borne disproportionately by low-income populations (*7*). This disparity, where those suffering most are often least responsible for the historical greenhouse gas (GHG) emissions causing the warming (*8*), is a fundamental global injustice demanding rigorous quantification and understanding.

Understanding the full scale of this disparity and attributing responsibility, however, remains hampered by methodological gaps. Previous global assessments focused primarily on mortality projections (*9–11*), often lacking fine spatial resolution or aggregating impacts, while comprehensive health burden assessments incorporating morbidity typically remained location-specific case studies (*12–20*), limiting global comparison and attribution and often excluding the most heavily impacted regions in Africa and Asia. An important complementary line of work is the Global Burden of Disease (GBD), which provides disaggregated, global estimates of the health burden attributable to non-optimal temperatures, reported in deaths and DALYs (*2*). However, GBD quantifies contemporaneous burden relative to a theoretical minimum risk exposure and does not attribute that burden to emitting producers or consumption in other regions, nor does it estimate time-integrated damage caused by a single year of GHG emissions. Critically, a globally consistent framework has been missing to directly link consumption-based GHG emissions to resulting health burdens. This is particularly true for emissions originating in high-income countries, and the temperature-related health impacts experienced differentially across specific populations (*8*, *21*).

Our methodological framework integrates data and models across three main stages. First, we build on recent advances in climate impact modeling (*11*) and global health data (*22*, *23*) to develop country-specific Damage Factors (DFs), which quantify the potential health burden (in DALYs) per degree of warming for 163 countries and regions. Second, we integrate these DFs with climate models of GHG fate and transport to derive Characterization Factors (CFs), capturing the health damages per unit of emission for major GHGs. We then apply these CFs to a global, consumption-based emissions inventory derived from a multi-regional input-output (MRIO) model (*24*). Finally, this allows us to attribute the total health damage induced by the consumption of each ‘inducer’ nation to the various ‘receiver’ nations, thereby quantifying the net health injustice.

### Unequal Global Health Burdens from Temperature Change

To quantify the health impacts of climate change, this study first establishes spatially differentiated Damage Factors (DFs) for 163 countries and regions (**Fig. 1**). These DFs represent the health burden from direct heat and cold exposure, in Disability-Adjusted Life Years (DALY), that is incurred per capita and per year for every cumulative degree Celsius-Year of global warming averaged over the entire globe. This metric integrates the health damage over time, reflecting that the temperature increase is a sustained state over centuries, not a single-year event. A higher DALY value per death in a given country, often resulting from higher mortality rates among younger populations, contributes to a higher overall damage factor, thus embedding demographic vulnerability directly into our analysis (**data S1–S3**).

**Fig. 1.**
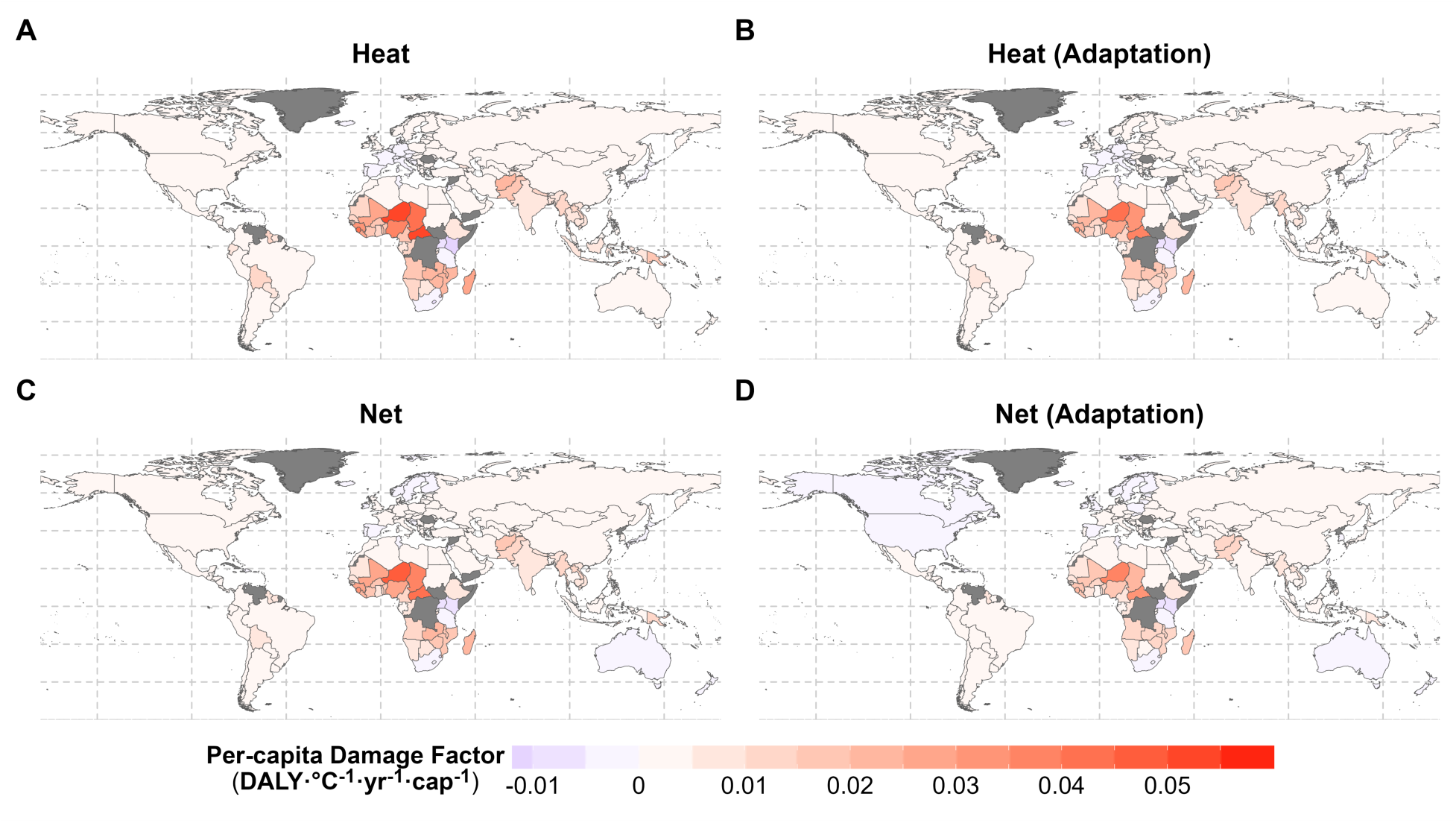
Unequal global health vulnerability to climate change. The maps show the per capita health Damage Factor (DF)—expressed in Disability-Adjusted Life Years (DALYs) per degree Celsius per year (DALY·°C^-1^·yr^-1^·cap^-1^) (**data S3**). Values represent an average across all Representative Concentration Pathway (RCP) scenarios and the two future time periods (2040–2059 and 2080–2099) considered in this study. (**A**) Damage Factor for heat. (**B**) Heat-related Damage Factor considering income-based adaptation measures (e.g., access to cooling). (**C**) The net Damage Factor from both heat and cold effects. (**D**) The net Damage Factor where adaptation measures are applied to the heat component. Red hues indicate higher vulnerability (net health burden), while blue hues indicate a net reduction in the temperature-related health burden, where reduced cold-related mortality outweighs increased heat damage. Data sources include mortality projections from Bressler et al. (2021) (**data S1**) and health burden data from the Global Burden of Disease 2019 study (**data S2**) (*11*, *22*, *23*). We quantified uncertainty in damage factors using Monte Carlo simulation (10,000 iterations) to derive 2.5^th^–97.5^th^ percentile intervals, with full results provided in Supplementary Materials and **data S4**.

Our initial analysis of net health damages, conducted for a baseline without modeled adaptation measures, reveals severe global inequalities rooted in both climatic exposure and socioeconomic vulnerability (**Fig. 1C and data S3**). The highest net health damage is overwhelmingly concentrated in lower-income and subtropical regions. Sub-Saharan Africa emerges as the most impacted continent, with nations like Niger (0.046 DALY·cap^-1^·°C^-1^·yr^-1^), the Central African Republic (0.045), and Chad (0.035) exhibiting the highest damage factors. This extreme vulnerability is associated with characteristics common to low-income nations, such as high baseline temperatures and economic constraints which hinder coping mechanisms. In these regions, increased mortality and morbidity from heat stress are the primary drivers of health damage, significantly outweighing any potential reductions in cold-related mortality (**Fig. 1A and data S3**).

Conversely, a small number of regions show net negative damage factors in this baseline scenario (**Fig. 1C**), especially high-altitude tropical countries like Kenya (-0.0094 DALY·°C^-1^·yr^-1^·cap^-1^) and high-latitude nations like Norway (-0.000428). This pattern is consistent with prior mortality-focused studies (*9*, *11*). These apparent net benefits are, however, modest—nearly five times smaller than the highest net damage factor observed in Niger. They reflect only the net balance of direct temperature-related DALYs and do not account for other severe climate impacts these regions face, including vector-borne disease burden and flood-related damages.

Incorporating income-based adaptation measures, representing societal capacity to implement protective strategies like cooling access and heat action plans, significantly mitigates heat-related damage universally (**Fig. 1B, 1D, and data S3**). For most nations facing a net health burden, this modeled adaptation leads to substantial reductions in that burden. For instance, Niger’s DF decreases from 0.046 to 0.035 DALY·°C^-1^·yr^-1^·cap^-1^ (∼24% reduction), and similar significant reductions are seen across many vulnerable nations. Conversely, for some high-income countries that already exhibit a small net benefit in the baseline scenario, adaptation can amplify this effect; Australia is a notable case where modeled adaptation turns a marginal net benefit into a much larger one. However, even with these shifts, profound inequalities persist. After adaptation is considered, a clear majority—representing 87.6% of the global population in our analysis—still face a net health damage. This underscores that adaptation, while essential, is insufficient on its own to counteract the escalating risks of heat stress globally, particularly in highly exposed and resource-limited regions. The remaining net health damage, primarily concentrated in low-income regions, is substantial, representing an annual loss of over 24 million healthy life years (DALYs) globally for every degree of sustained warming (**Fig. 1D and data S3**).

### Socioeconomic Factors Dominate Vulnerability

National capacity to cope with these temperature changes is overwhelmingly linked to socioeconomic development (represented by GDP per capita), largely independent of the specific future emissions trajectory within the studied range (**Fig. 2**). Our analysis across multiple RCPs (2.6 to 8.5) and future time periods reveals per-capita income as the primary determinant of projected health burdens per degree of warming (DALY·°C^-1^·yr^-1^·cap^-1^). Notably, the overlapping boxplots in **Fig. 2** illustrate that the choice of emission scenarios has no significant impact on the central estimate of vulnerability per °C-yr of the average temperature increase for a given country. This strongly suggests that near-to-mid-century vulnerability patterns are largely locked in by existing socioeconomic conditions and baseline health status, factors not substantially altered by different climate mitigation pathways over these timescales in the model framework.

**Fig. 2.**
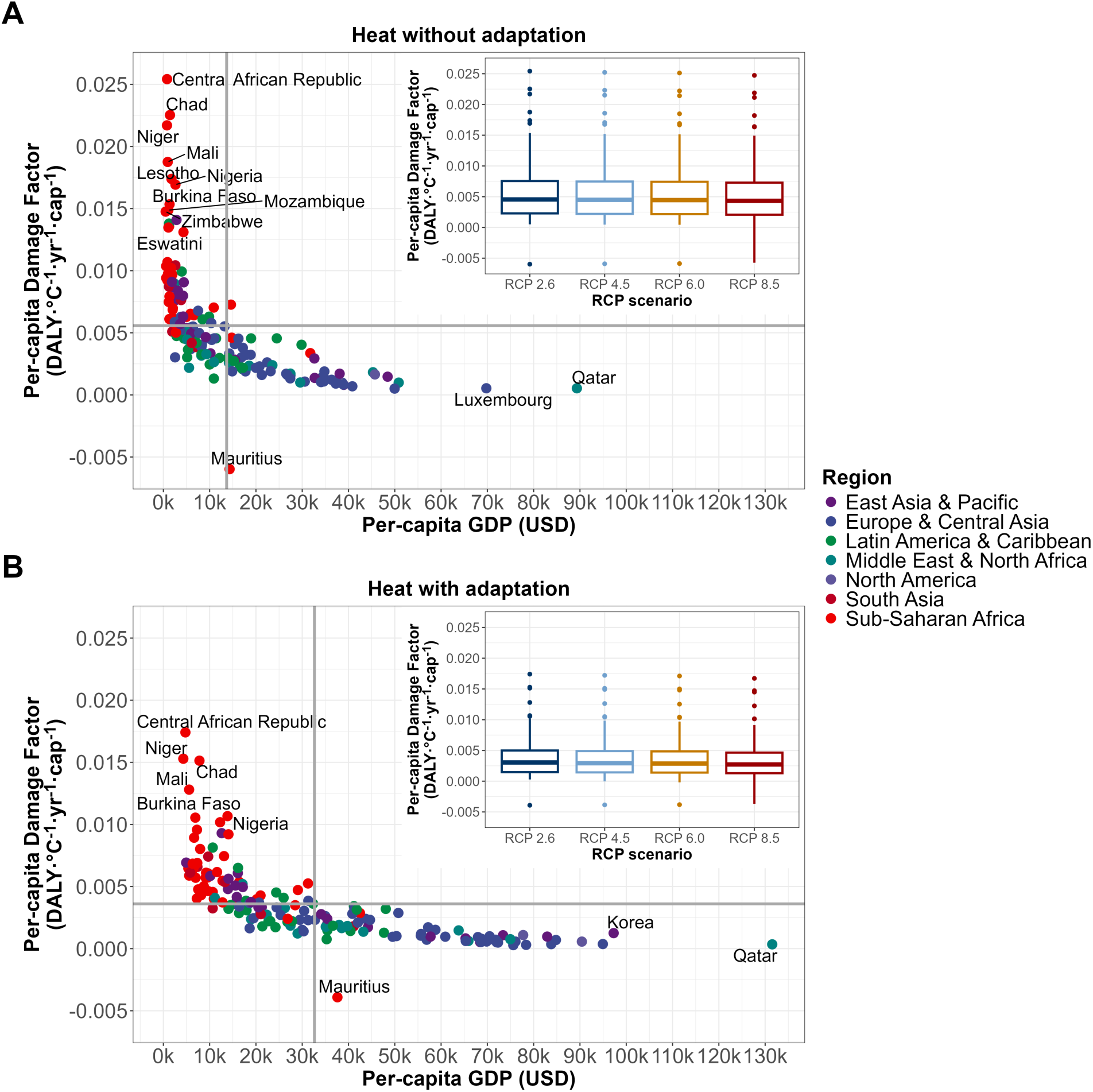
Socioeconomic development overwhelmingly determines health vulnerability to temperature-related climate impacts. The per capita health Damage Factor (DF) (DALY·°C^-1^·yr^-1^·cap^-1^) is plotted against per capita Gross Domestic Product (GDP, in USD) for 163 countries and regions. For visual clarity, the scatter plot points represent a country under the optimistic Representative Concentration Pathway (RCP) 2.6 scenario for the future time period of 2080–2099; colors indicate geographical regions. (**A**) Relationship for heat-related DFs without adaptation. (**B**) Relationship for heat-related DFs including adaptation measures linked to future GDP projections. Inset boxplots show the full distribution of the Damage Factor across all four RCP scenarios, highlighting the limited influence of the emissions scenario compared to national income. Horizontal and vertical grey lines indicate the mean values. All box plots comprise the median line, the box indicates the interquartile range (IQR), whiskers denote the rest of the data distribution and outliers are denoted by points greater than ±1.5 × IQR. Details are provided in the Supplementary Materials and **data S3**.

A strong, non-linear inverse relationship exists between per capita income and the health damage factor (**Fig. 2 scatter plot and data S3**). Vulnerability decreases sharply with rising GDP, eventually clustering near zero or becoming negative (reflecting the cold mortality effect) for countries with GDP exceeding roughly $30,000 per capita. This indicates that GDP per capita serves as a robust proxy for a wide array of adaptive capacities: access to resilient energy and building infrastructure, advanced healthcare and public health systems, effective governance including early warning systems, and individual resources for protective behaviors (*1*). Consequently, low-income nations, primarily located in Sub-Saharan Africa and parts of South Asia, consistently demonstrate the highest vulnerability (with several nations exceeding 0.02 DALY·°C^-1^·yr^-1^·cap^-1^), regardless of the climate scenario considered. Projecting future economic growth alongside adaptation measures (**Fig. 2B**) reinforces this fundamental relationship: while overall vulnerability may decrease slightly as low-income countries develop, the curve retains its shape, emphasizing that higher income strongly correlates with greater resilience to direct temperature impacts. This highlights potential health co-benefits of sustainable development pathways but also the deep-seated nature of current climate vulnerability.

### Consumption Drives Global Health Injustice

To connect health impacts to their underlying drivers, we derived global average characterization factors (CFs) quantifying the long-term health burden (DALYs) per kilogram of GHG emitted, integrating atmospheric science, climate modeling, and epidemiological response, while incorporating adaptation potential (**Table 1**, **data S5, and data S6**). These CFs vary dramatically between GHGs due to distinct radiative efficiencies and atmospheric lifetimes. Calculated over a 500-year time horizon, the health impact per kilogram of fossil methane (CH_4_: 4.52E-05 DALY·kg^-1^ with adaptation) or long-lived nitrous oxide (N_2_O: 5.85E-04 DALY·kg^-1^) is thus 1-2 orders of magnitude greater than that of the reference gas, carbon dioxide (CO_2_: 4.48E-06 DALY·kg^-1^) (**Table 1 and data S5**). Highly potent industrial gases like sulphur hexafluoride (SF_6_: 1.52E-01 DALY·kg^-1^) exhibit extremely high CFs, emphasizing the importance of controlling their emission sources despite lower emission volumes compared to CO_2_.

**Table 1.** Global average human health characterization factors (CFs) for the major greenhouse gases covered by the Kyoto Protocol and IPCC AR6. CFs quantify the long-term human health damage over a 500-year time horizon per kilogram of greenhouse gas (GHG) emitted, expressed in DALY per kilogram (DALY·kg ^-1^). Factors are presented considering societal adaptation measures to net impacts of climate change due to heat and cold. These global average factors integrate the atmospheric fate of GHGs, their contribution to temperature increase, and the resulting impacts on mortality due to heat and cold (**data S5**). Data are shown for 10 major GHGs covered by the Kyoto Protocol or presented in Table 7.15 of IPCC AR6; a full list is available in **data S5**. These CFs are used to translate GHG emissions into health outcomes.

| Substance | CF net adjusted (Deterministic) | CF net adjusted (Monte Carlo, 95% CI) |
| --- | --- | --- |
| CO<sub>2</sub>*† | 4.48·10<sup>-6</sup> | 4.89·10<sup>-6</sup> (1.97·10<sup>-6</sup>–8.74·10<sup>-6</sup>) |
| CH<sub>4</sub>* | 4.48·10<sup>-5</sup> | 5.13·10<sup>-5</sup> (1.21·10<sup>-5</sup>–1.20·10<sup>-4</sup>) |
| N<sub>2</sub>O* | 5.82·10<sup>-4</sup> | 6.71·10<sup>-4</sup> (1.57·10<sup>-4</sup>–1.58·10<sup>-3</sup>) |
| CFC-11 | 9.38·10<sup>-3</sup> | 1.07·10<sup>-2</sup> (2.84·10<sup>-3</sup>–2.41·10<sup>-2</sup>) |
| PFC-14 | 4.74·10<sup>-2</sup> | 5.40·10<sup>-2</sup> (1.64·10<sup>-2</sup>–1.17·10<sup>-1</sup>) |
| HFC-134a | 1.95·10<sup>-3</sup> | 2.21·10<sup>-3</sup> (6.07·10<sup>-4</sup>–4.95·10<sup>-3</sup>) |
| HFC-32 | 9.86·10<sup>-4</sup> | 1.12·10<sup>-3</sup> (3.06·10<sup>-4</sup>–2.52·10<sup>-3</sup>) |
| SF<sub>6</sub>* | 1.53·10<sup>-1</sup> | 1.77·10<sup>-1</sup> (4.22·10<sup>-2</sup>–4.13·10<sup>-1</sup>) |
| NF<sub>3</sub>* | 8.15·10<sup>-2</sup> | 9.37·10<sup>-2</sup> (2.60·10<sup>-2</sup>–2.11·10<sup>-1</sup>) |

\*Used for the consumption based analysis.
†Also used for average PFCs whose inventories are directly reported in kg CO<sub>2</sub> equivalent.

Applying these CFs to national consumption-based GHG inventories (derived using MRIO model Eora and PRIMAP-hist data (*24*, *25*); **data S6**) allows tracing the health consequences induced by consumption and resulting economic activity, revealing a quantifiable global climate health injustice (**Fig. 3 and data S6**). A dramatic geographical mismatch and inverse correlation exists between where consumption-driven emissions effectively originate and where the resulting health burdens manifest. High-income regions (North America, Europe & Central Asia comprising less than 20% of the global population) induce together with East-Asia & Pacific the vast majority of consumption-related health damage due to their high per-capita consumption and associated trade-related emissions. The per capita damage induced by North American consumption reaches 0.09 DALY·cap^-1^·year^-1^ (**Fig. 3A**), a figure equivalent to imposing an average loss of 34 days of healthy life per person, per year, on the global population.

**Fig. 3.**
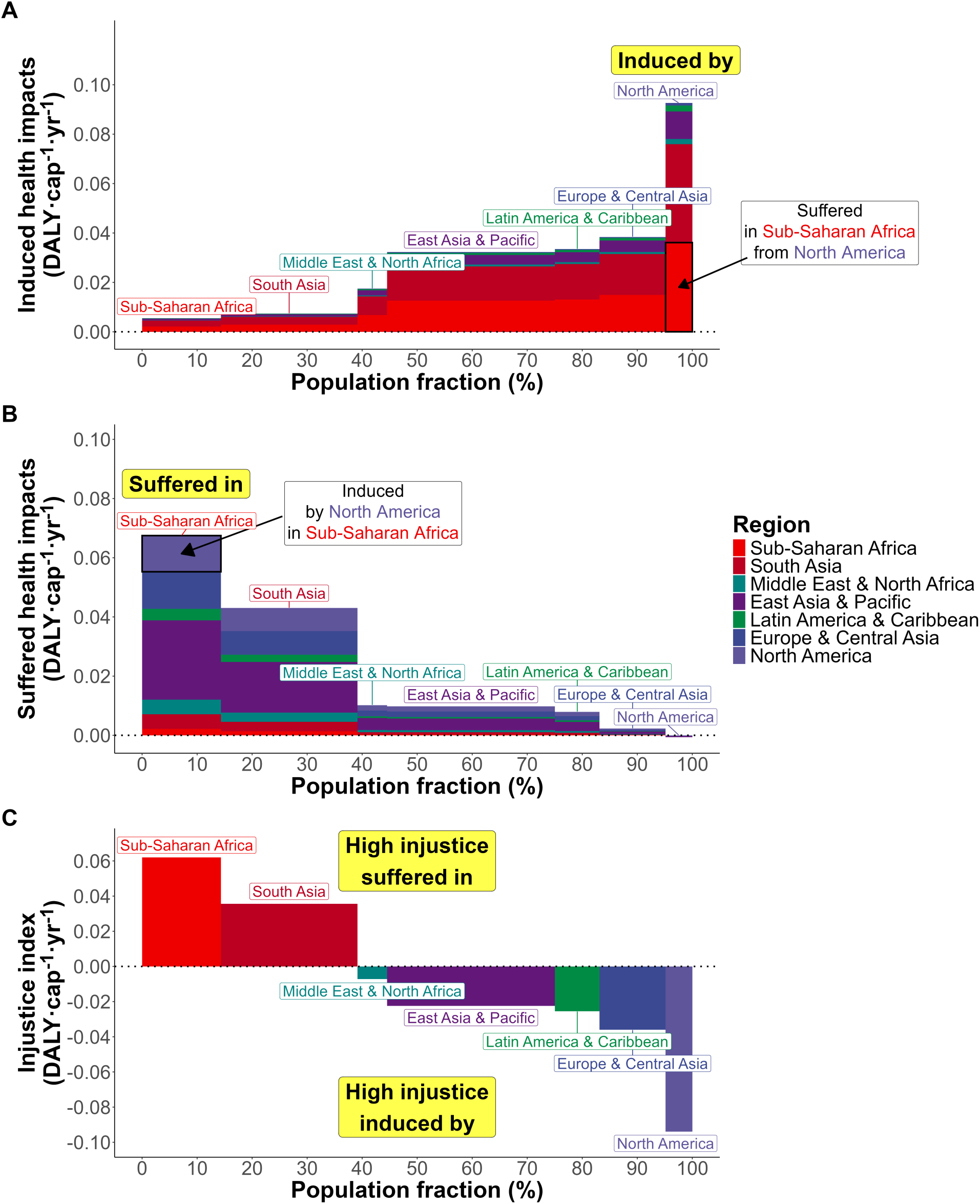
Consumption in high-income regions drives the global climate health burden. The figure displays the per capita Health Damage (DALY·cap^-1^·yr^-1^) from climate change, attributed to annual consumption-based emissions (**data S6**). Regional totals are aggregated from 163 individual countries and territories. The width of each regional block along the x-axis is proportional to its share of the global population, meaning the area of each rectangle represents the total health damage for that segment. (**A**) The ‘Inducer’ perspective, showing the per capita health damage caused globally by each region’s consumption patterns. (**B**) The ‘Receiver’ perspective, showing the per capita health damage suffered by each region, irrespective of who caused the emissions. (**C**) The ‘Health Injustice Index’, calculated as received damage minus induced damage (B - A). Positive values indicate regions that suffer a disproportionately high burden (net sufferers), while negative values indicate regions that are net sources of health damage globally (net inducers). All calculations incorporate adaptation effects.

In clear contrast stand the populations suffering the greatest health burden per capita from climate change, irrespective of their own emission levels. Sub-Saharan Africa experiences the highest received burden at approximately 0.07 DALY·cap^-1^·year^-1^ (∼25 days/year), with South Asia also bearing a very high burden (**Fig. 3B**). These regions endure this disproportionate suffering despite their minimal contribution to the global consumption patterns driving climate change. This illustrates the externalization of health costs from high-consumption lifestyles, predominantly in the Global North, onto vulnerable populations, predominantly in the Global South. Intriguingly, some high-income regions (notably North America) show slightly negative received burdens per capita on average. This is a net effect of regional aggregation, in which the reduction of cold-related burdens in high-latitude countries (e.g., USA and Canada) slightly outweighs the heat-related damage experienced in lower-latitude areas (e.g., Mexico) when averaged across the entire region’s population.

We define the Health Injustice Index to quantify this imbalance; it is expressed as the received minus induced damage per capita. When computing this metric for the different regions (**Fig. 3C**), a clear division emerges between net sufferers and net inducers of climate health damage. Sub-Saharan Africa displays the largest net positive index (+0.062 DALY·cap^-1^·year^-1^), representing a net health burden equivalent to approximately 23 lost days of healthy life per person per year. South Asia also emerges as a significant net sufferer (+0.036 DALY·cap^-1^·year^-1^). These positive values signify that the health damage these regions suffer from the consumption of other nations is greater than the damage they export externally. Conversely, all other regions function as net inducers, with North America showing the largest net negative index (-0.094 DALY·cap^-1^·year^-1^), equivalent to a net export of health harm of roughly 34 days per person per year. Europe & Central Asia, Latin America & Caribbean, and East Asia & Pacific also function as significant net inducers (**Fig. 3C**). The global map of this net injustice (**Fig. 4**) vividly portrays this dynamic: industrialized and high-consuming nations, predominantly shaded blue, externalize severe health burdens onto vulnerable, low-emitting nations, predominantly shaded red, creating a clear geography of climate health injustice.

**Fig. 4.**
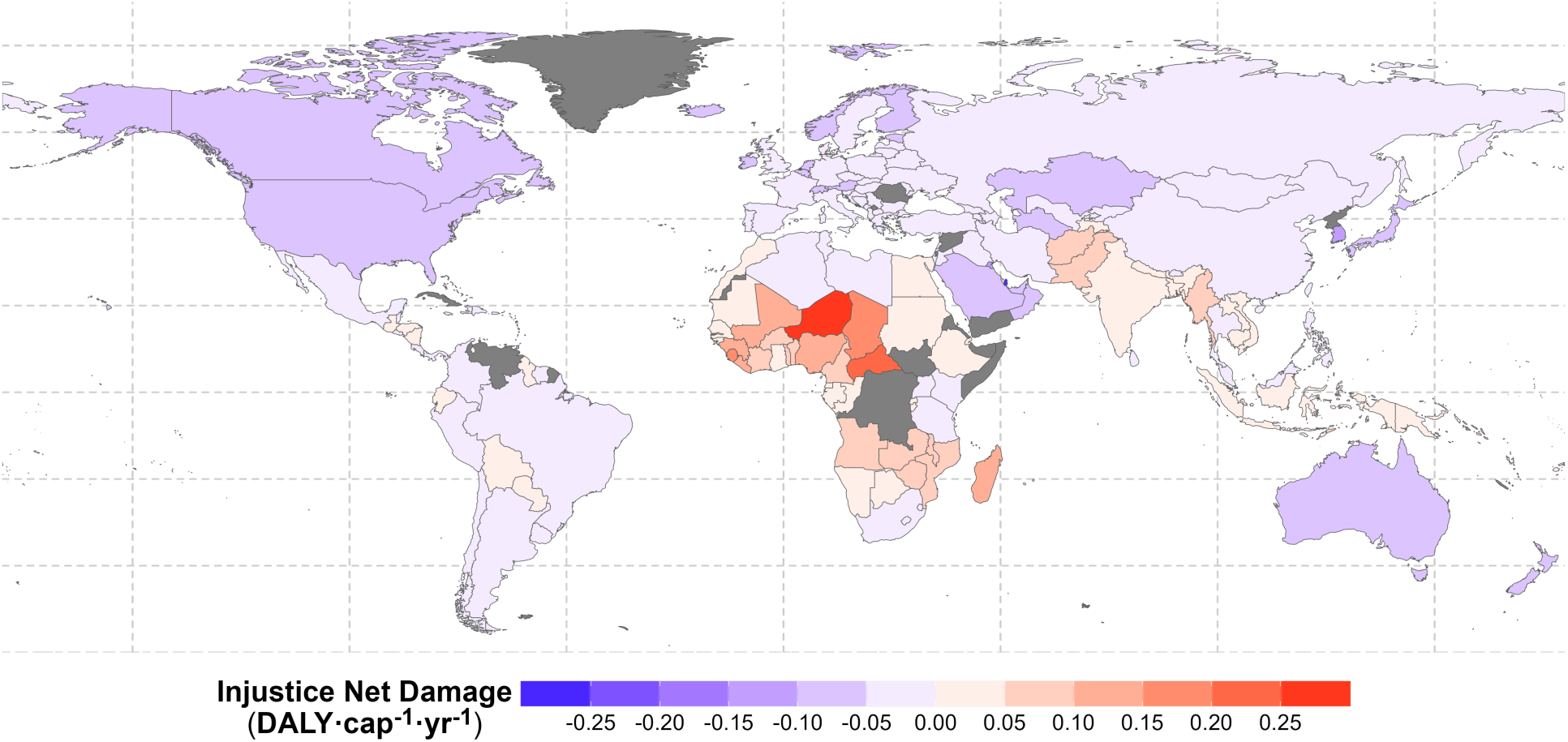
The country-specific global geography of consumption-driven climate health injustice. The map displays the per capita Health Injustice Index (DALY·cap^-1^·yr^-1^), calculated as the health damage received by a country minus the health damaged induced by its consumption-based GHG emissions (Received - Induced) (**data S7**). Red/orange hues indicate countries and regions with a positive index, representing net sufferers who endure disproportionately high health damage relative to their own consumption. Blue/purple hues indicate countries and regions with a negative index, representing net inducers whose consumption drives health damage elsewhere. Grey indicates no data or excluded regions. All calculations incorporate adaptation effects.

### Implications for Climate Equity and Policy

Our analysis provides a quantitative framework that directly links national consumption patterns to spatially explicit global health burdens from climate change, offering a powerful, health-focused lens for understanding and addressing climate injustice. By translating GHG emissions into regionally specific DALYs, we map the unequal flow of climate-related harm embedded within the structure of the global economy. The quantification demonstrates that consumption imposes measurable health costs, equivalent potentially to months of lost healthy life per year, on populations least responsible. It underscores the profound ethical dimensions and inequities of the climate crisis. This challenges narratives focused solely on territorial emissions and highlights the far-reaching global health responsibilities associated with high-consumption lifestyles.

These findings hold critical implications for international climate policy and the pursuit of global equity. The demonstrated dominance of socioeconomic factors in determining near-term vulnerability (**Fig. 2**) reinforces the urgent need for significantly scaled-up, proactive, and targeted adaptation finance and support flowing to low-income nations. Investments must prioritize strengthening primary health care systems, building climate-resilient infrastructure (water, energy, housing), enhancing social safety nets, and implementing effective early warning systems to reduce baseline vulnerability before impacts escalate further. Furthermore, our consumption-based attribution of health burdens (**Figs. 3 and 4**) provides a crucial health-centric evidence base for negotiations regarding equitable mitigation contributions under the Paris Agreement (Nationally Determined Contributions, NDCs), the international climate finance architecture—including the Green Climate Fund, the Adaptation Fund, and the recently operationalized fund for Loss and Damage (*26*)—and accountability frameworks. It offers concrete, health-based data to inform discussions on common but differentiated responsibilities and respective capabilities, moving beyond purely economic or emissions-based metrics.

This DALY-based quantification of health injustice complements other important approaches like the Social Cost of Carbon (SCC) (*27*). While SCC provides vital aggregated monetary estimates, our bottom-up, health-focused methodology translates complex climate impacts into a universally understood metric of human well-being—lost healthy life years. A key finding is the relative stability of the per-degree Damage Factors across different emissions scenarios (**Fig. 2**), suggesting these factors can serve as a robust tool for future policy assessments. While our analysis focuses on temperature-related direct impacts (see Supplementary Materials for a detailed discussion of scope and uncertainties) and accounts for both substantial reductions from income-based adaptation and localized offsets from reduced cold mortality (*9*, *11*), our comprehensive mapping of the injustice dynamics (**Figs. 3 and 4**) demonstrates that these combined mitigating effects are vastly outweighed by the immense health burdens imposed elsewhere, which affect a clear majority of countries representing nearly 88% of the global population in our analysis.

This work provides a novel, health-centric evidence base for climate accountability. By offering a tool to characterize and monitor the health impacts of consumption, it can directly inform mechanisms like the Loss and Damage fund where accountability is paramount. Ultimately, this quantification creates a pathway toward policies where responsibility is measured not just in tonnes of carbon, but in the tangible costs imposed on human health. While this study focuses on temperature-related burdens, our framework provides a scalable foundation designed to integrate other human health impacts of climate change as epidemiological data become available, creating a more holistic picture of our global responsibilities in this warming world.

## Acknowledgments

## Funding

Novo Nordisk Foundation grant NNF22OC0075778 (DY, OJ)

Fraunhofer-DTU project EDES (eco-design Stewardship), advancing eco-design excellence, supporting the aeronautics European – Clean Sky Programme (LR, ALaurent)

Canada Research Chair on Measuring the Impact of Human Activities on Climate Change (ALevasseur, CA)

## Author contributions

Conceptualization: LR, OJ, DY

Methodology: LR, OJ, ALevasseur, CA

Visualization: LR, OJ, DY

Funding acquisition: ALevasseur, ALaurent, OJ

Project administration: ALaurent, OJ

Supervision: ALaurent, OJ

Writing – original draft: LR, DY

Writing – review & editing: LR, DY, ALevasseur, ALaurent, OJ

## Data and materials availability

Complete code and data to reproduce the figures are available in the public GitHub repository: https://github.com/udaniel/climate-health-injustice. All data are available in the main text or the supplementary materials.

