## Supplementary Methods for "Global Health Injustice From Climate Change Driven By Consumption"

†Equal contribution

### Conceptual framework to assess damage to human health associated with consumption

Consumption in inducing country *i* generates GHG emissions that lead to a time integrated increase in global mean surface temperature. This temperature rise triggers climate-related events such as heatwaves and cold waves, which cause death in the receiving country *c* and therefore a human health loss in that country expressed in Disability-adjusted life years (DALY), see Fig. S1.

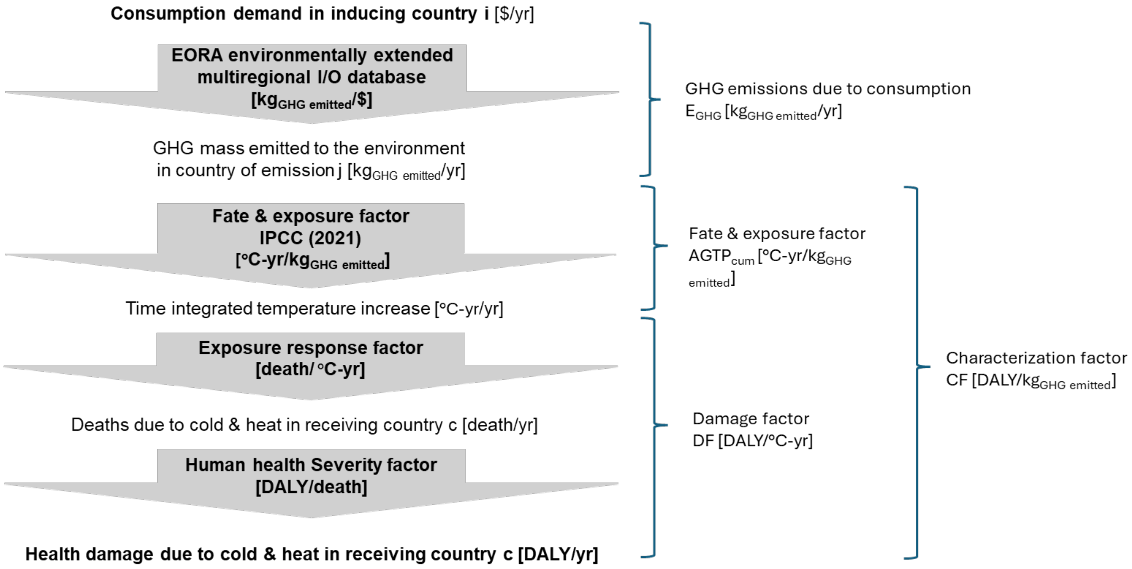


**Fig. S1.** Visual representation of the impact pathway for damage to human health due to cold and heat, induced by the total consumption. The impact framework uses a global environmentally extended database to first determine the GHG emissions induced by the total annual consumption ($E_{GHG}$). These emissions are then multiplied by the fate and exposure factor ($AGTP_{cum}$) to obtain the cumulative temperature increase. We then developed the damage factor (DF) as the combination of a temperature exposure response factor and a severity factor to finally yield the resulting health damage due to heat and cold (D). Abbreviations: DALY, disability-adjusted life years; GHG, greenhouse gas.

The total annual health damage for health risk ***r*** (heat/cold/net) in receiving country ***c*** ($D_{r,i \rightarrow c}$), due to the consumption-based emissions of inducing country $i$, is expressed in disability-adjusted life years (DALY) per year [DALY· $yr^{-1}$] and calculated according to Equation S1 (see also Fig. S1):

$$D_{r,i\rightarrow c} \;=\; \sum_{k}\sum_{h}\left(DF_{\mathrm{c,r,h}} \times AGTPcum_{k,h}\right) \;\times\; E_{GHG\;k,i} = \sum_{k} CF_{\mathrm{c,r,k}} \;\times\; E_{GHG\;k,i}$$

(Equation S1)

We first multiply the damage factors specific to receiving country ***c*** , health risk ***r***, and time horizon period ***h*** ($DF_{\mathrm{c,r,h}}$, in DALY·°C$^{-1}$·yr$^{-1}$), by the Cumulative Absolute Global Temperature Change Potential per kg GHG ***k*** over time horizon period ***h*** ($AGTP_{cum,k,h}$, in °C·yr·$kg_{GHG}^{-1}$) and sum up over the two time horizon periods h (0-100 and 100-500 years) to yield the characterization factor representative of the health damage for risk ***r*** in receiving country ***c*** per $kg_{GHG}$ k emitted ($CF_{c,r,k}$ in DALY·$kg_{GHG}^{-1}$). The total annual health damage in country $c$ induced by consumption in country $i$ is then calculated by multiplying this characterization factor by the annual GHG emission induced by the total consumption in inducing country $i$ ($E_{GHG\;k,i}$, in $kg_{GHG}$·yr$^{-1}$).

The spatially differentiated Damage Factors (DF) for receiving country c quantifies the health damage per unit cumulative temperature change due to heat and cold from climate change, expressed in DALY·°C$^{-1}$·yr$^{-1}$, over two time periods, 0 to100 and 100 to 500 years. Its calculation is detailed in section S2 below.

$AGTP_{cum\;k,h}$ represents the cumulative temperature increase over time period $h$ due to the emission of GHG $k$ expressed in [°C·yr·kg $_{GHG}^{-1}$] and the characterization factor ($CF_{c,r,k}, in$ DALY·$kg_{GHG}^{-1}$) links the mass of GHG $k$ released to the final human health damage per kg $_{GHG}$ emitted over a time horizon period $h$. Their calculations are detailed in section S3 below.

The annual GHG emissions induced by the total consumption in inducing region $i$ ($E_{GHG\;k,i}$, in $kg_{GHG}$·yr$^{-1}$) is determined using the EORA global input-output model (*24*). Their calculations are detailed in section S4 below.

**Damage factor: Human health damage due to temperature change (heat/cold/net)**

We first calculate spatially differentiated damage factors for each receiving country c, health risk r, climate scenario $s_p$, and time period p, $DF_{c,r,s_p}$, using the Bressler et al. framework for cold, heat, heat with adaptation, net, and net with adaptation (*11*). Equation S2 presents the computation of these annual damage factors $DF_{c,r,s_p}$, expressed in [DALY·°C$^{-1}$·yr$^{-1}$], as a combination of 1) the temperature-attributable mortality relative risk $RR_{c,r,s_p}$ [%] from Bressler et al. (*11*), 2) the total annual deaths $TD_c$, [death] in receiving country *c* and the DALY per death $DD_{c,r}$ [DALY·death$^{-1}$] in country *c* due to risk *r* (heat or cold) taken from the Global Burden of Disease (GBD) for the year 2019, and 3) the projected change in global mean surface temperature $\Delta T_{g,s_p}$ [°C] between the reference 1986-2005 period and the two mid and late 21st century periods p (2040–2059 and 2080–2099) (*22*, *23*), for the four RCP scenarios $s_p$ (RCP2.6, RCP4.5, RCP6.0, and RCP8.5) applied to these two periods p:

$$DF_{c,r,s_p} = \frac{RR_{c,r,s_p} \times TD_c \times DD_{c,r}}{\Delta T_{g,s_p}}$$

(Equation S2a)

The net damage factor is then calculated as the sum of the heat and cold damage factors.

$$DF_{c,net,s_p} = DF_{c,heat,s_p} + DF_{c,cold,s_p}$$

(Equation S2b)

Further description of the parameters used in Equation S2 is provided in the following sections.

*Relative risks for heat and cold (RR)*

Relative risk factors quantify the temperature-attributable percent change in mortality due to heat or cold as a function of the annual average temperature change at the country level resulting from climate change. Here, RR denotes the percent change in mortality (not a risk ratio), and we retain the original notation for consistency. The minimal mortality temperature for each location was determined to compute excess mortality attributable to temperature variations. Deviations from the minimal mortality temperature were associated with mortality risks due to heat or cold, respectively (*2*).

The most extensive global extrapolation of excess mortality, covering 166 countries for both heat and cold, was conducted by Bressler et al. (*11*). This model was used to compute damage factors per country under different scenarios: heat, cold, heat with adaptation, net, and net with adaptation. We adopted the approach from Bressler et al. (*11*), which defines relative risks (*RR)* separately for heat and cold, combining the non-linear effects of warming per country, Gross

Domestic Product Per Capita $\log(GDPPC)_{c,s_p}$ under scenarios (e.g., Shared Socioeconomic Pathway (SSP) projections), as well as prevailing climate conditions. The equations for heat- and cold-related risks are given below as Equations S3 and S4, respectively.

$$RR_{c,heat,s_p} = \beta_1 + \beta_2 \times \Delta T_{c,s_p} + \beta_3 \times \bar{T}^{mmax}_{c,2020} + \beta_4 \times \log(GDPPC)_{c,s_p}$$

(Equation S3)

$$RR_{c,cold,s_p} = \beta_1 + \beta_2 \times \Delta T_{c,s_p} + \beta_3 \times \bar{T}^{mmin}_{c,2020}$$

(Equation S4)

where *c* represents the country, *s* denotes the RCP scenario, and *p* refers to the time period within the RCP scenario periods. $RR_{c,heat,s_p}$ represents the percentage change in mortality due to heat, while $RR_{c,cold,s_p}$ represents the same for cold. The coefficients $\beta_1, \beta_2, \beta_3$, and $\beta_4$ are fixed baseline covariates that describe prevailing climate conditions and are taken directly from Bressler et al. (2021) (see Table S1 and data S1). $\bar{T}^{mmin}_{c,2020}$ and $\bar{T}^{mmax}_{c,2020}$ are the average temperatures for the hottest and coldest months in country c calculated as population-weighted averages. The $\log(GDPPC)_{c,s_p}$ represents the projected Gross Domestic Product per capita for country *c* under the given RCP scenario (*21*). The country-specific temperature change $\Delta T_{c,s_p}$ for each RCP scenario and period p was derived as described in Section 2.5.

*Total death (TD) and DALY/death (DD)*

The annual number of deaths in each country ($TD_c$) represents the total annual death due to all-cause mortality, obtained from the Global Burden of Disease (GBD) dataset for the year 2019 and these values are held constant across scenarios (*22*, *23*). Given the variation in underlying causes of death—such as age, risk factors, and healthcare system differences, we introduce the parameter DALY per death ($DD_{c,r}$), which quantifies the average DALYs per death in country $c$ for health risk $r$ associated with temperature change. For this parameter, we use country specific data on non-optimal temperature-related health risks from GBD 2019 (*22*, *23*). Accordingly, $RR_{c,r,s_p}$ is applied to all-cause baseline mortality while $DD_{c,r}$, scales deaths into DALYs under temperature-related burden assumptions. In this study, low temperatures in GBD serve as a proxy for cold-related damage, while high temperatures correspond to heat-related damage. Each risk category in GBD is linked to a range of diseases contributing to mortality and morbidity, including communicable and non-communicable diseases. Detailed country-level values for $TD_c$ and $DD_{c,r}$ are provided in data S2.

*Climate scenarios*

The spatially differentiated damage factors for heat- and cold-related health impacts were developed based on the modelling scenarios from Bressler et al. 2021 (*11*). For each country, four Representative Concentration Pathways (RCPs)—RCP2.6, RCP4.5, RCP6.0, and RCP8.5—are considered across two time periods (2040–2059 and 2080–2099). Additionally, an adaptation scenario, modeled as a function of income, is available exclusively for heat-related outcomes. This simplified approach assumes that higher income levels can mitigate heat-related damage, for example, through increased access to air conditioning.

RCP scenarios originate from the Intergovernmental Panel on Climate Change (IPCC) Fifth Assessment Report (AR5) in 2014 and represent different climate futures based on varying levels of GHG emissions over time (*28*). The numbering of each scenario corresponds to the projected radiative forcing values in $W/m^2$ by the year 2100. We retain these AR5 RCPs to remain consistent with the Bressler et al. 2021 modelling framework (*11*).

Results for all damage factors—stratified by country, RCP scenario, time period, and cause-specific damage (heat, cold, heat with adaptation, net, and net with adaptation) are available in data S3. Net denotes the combined heat and cold effect; net with adaptation applies income-based adaptation to the heat component only. Furthermore, we introduce shorter (100-year) and longer (500-year) time horizons to reflect the different atmospheric lifetimes of greenhouse gases, which range from a few years to several thousand years, and report the resulting aggregated damage factors (DFs).

*Aggregation of RCP scenarios to obtain DFs for main horizon periods*

Damage factor are spatially differentiated for 166 countries and as the per degree results are largely independent of the temperature increase and of the considered RCP scenarios (figure 2 main text), they can be averaged across all scenario–period combinations reported by Bressler et al. (2021) to obtain best estimates of DF value for short-term (100yr) and long-term (500yr) CF calculations

(*11*). For the h=0-100 year horizon period, DFs were derived by averaging results for the two available future periods (p2040-2059 and 2080-2099) across all RCP scenarios. For the h=0-500 year horizon period, long-term impacts were approximated using the late-century period (2080-2099) across all RCP scenarios, consistent with the available modelling outputs:

$$DF_{c,r,0-100} = \frac{1}{8}\sum_{s_p}\left(DF_{c,r,s_{2050-2059}} + DF_{c,r,s_{2080-2099}}\right) \qquad \text{(Equation S5)}$$

$$DF_{c,r,100-500} = \frac{1}{4}\sum_{s_{2080-2099}}\left(DF_{c,r,s_{2080-2099}}\right) \qquad \text{(Equation S6)}$$

Here, $DF_{s_p}$ denotes the damage factor for a given scenario–period combination $s_p$, where $p \in$ {2050-2059, 2080-2099} and $s \in$ {RCP 2.6, 4.5, 6.0, 8.5}. While calculations are first made separately for each of the countries considered (data S3) to be able to identify which countries are most or least impacted, these damages factors can also be summed up across all countries to yield a global aggregated damage factor for each RCP scenario and as a total (Table S1). Since damage factors are determined per °C, the variability of their values is limited across scenarios. Deterministic and probabilistic averages are close, within 10% of each other. As expected, the uncertainty is lower for the independent scenario (lower ±17% 95$^{th}$ CI) than for the upper end dependent scenario (up to ±76% 95$^{th}$ CI). Confidence intervals are mostly symmetrical, DF being close to normal distribution.

**Table S1.** Global aggregated absolute damage factors per RCP scenario and cause (cold, heat, heat with adaptation, net, and net with adaptation) for short- and longer-term periods. These values represent the total global health burden (summed across all populations) and are used to derive the final global Characterization Factors. Comparison between deterministic and probabilistic global damage factors with uncertainties.

| Climate Scenario | Year | $DF_{cold}$ | $DF_{heat}$ | $DF_{heat}$ with adaptation | $DF_{net}$ | $DF_{net}$ with adaptation |
|---|---|---|---|---|---|---|
| | | [DALY·°C$^{-1}$·yr$^{-1}$] | [DALY·°C$^{-1}$·yr$^{-1}$] | [DALY·°C$^{-1}$·yr$^{-1}$] | [DALY·°C$^{-1}$·yr$^{-1}$] | [DALY·°C$^{-1}$·yr$^{-1}$] |
| RCP 2.6 | 2040-2059 | $-9.52 \cdot 10^{6}$ | $4.11 \cdot 10^{7}$ | $3.26 \cdot 10^{7}$ | $3.16 \cdot 10^{7}$ | $2.31 \cdot 10^{7}$ |
| RCP 2.6 | 2080-2099 | $-7.06 \cdot 10^{6}$ | $3.29 \cdot 10^{7}$ | $2.18 \cdot 10^{7}$ | $2.58 \cdot 10^{7}$ | $1.48 \cdot 10^{7}$ |
| RCP 4.5 | 2040-2059 | $-1.32 \cdot 10^{7}$ | $4.92 \cdot 10^{7}$ | $3.86 \cdot 10^{7}$ | $3.60 \cdot 10^{7}$ | $2.54 \cdot 10^{7}$ |
| RCP 4.5 | 2080-2099 | $-1.60 \cdot 10^{7}$ | $6.10 \cdot 10^{7}$ | $4.01 \cdot 10^{7}$ | $4.50 \cdot 10^{7}$ | $2.41 \cdot 10^{7}$ |
| RCP 6.0 | 2040-2059 | $-1.02 \cdot 10^{7}$ | $4.01 \cdot 10^{7}$ | $3.18 \cdot 10^{7}$ | $2.99 \cdot 10^{7}$ | $2.16 \cdot 10^{7}$ |
| RCP 6.0 | 2080-2099 | $-1.54 \cdot 10^{7}$ | $6.16 \cdot 10^{7}$ | $4.04 \cdot 10^{7}$ | $4.62 \cdot 10^{7}$ | $2.50 \cdot 10^{7}$ |
| RCP 8.5 | 2040-2059 | $-1.28 \cdot 10^{7}$ | $4.82 \cdot 10^{7}$ | $3.77 \cdot 10^{7}$ | $3.54 \cdot 10^{7}$ | $2.49 \cdot 10^{7}$ |
| RCP 8.5 | 2080-2099 | $-1.16 \cdot 10^{7}$ | $6.06 \cdot 10^{7}$ | $3.91 \cdot 10^{7}$ | $4.90 \cdot 10^{7}$ | $2.75 \cdot 10^{7}$ |
| **Aggregated net DF (0-100-year) deterministic** | | | | | **$3.74 \cdot 10^{7}$** | **$2.33 \cdot 10^{7}$** |
| **Aggregated net DF (0-100-year) average probabilistic mean* (95th CI)** | | | | | **$3.93 \cdot 10^{7}$ ($2.28 \cdot 10^{7}$ - $5.61 \cdot 10^{7}$)** | **$2.47 \cdot 10^{7}$ ($1.35 \cdot 10^{7}$ - $3.61 \cdot 10^{7}$)** |
| **Aggregated net DF (100-500-year) deterministic** | | | | | **$4.15 \cdot 10^{7}$** | **$2.29 \cdot 10^{7}$** |
| **Aggregated net DF (100-500-year) average probabilistic mean* (95th CI)** | | | | | **$4.53 \cdot 10^{7}$ ($2.57 \cdot 10^{7}$ - $6.54 \cdot 10^{7}$)** | **$2.51 \cdot 10^{7}$ ($1.35 \cdot 10^{7}$ - $3.69 \cdot 10^{7}$)** |

*Medians are virtually equal to means with differences lower than 1% for all DFs

*Global temperature increase*

To ensure consistency with Bressler et al. (2021) (*11*), the projected change in global mean surface temperature for the mid and late 21st century, relative to the 1986-2005 baseline, is provided by the IPCC, AR5 (see Table S2). To align these projections with the baseline convention used by Bressler et al. (2021) (*11*), we computed the global temperature change $\Delta T_{g,s_p}$ by subtracting the observed warming in 2010 (0.72°C) from the IPCC projected values. Country-specific temperature changes $\Delta T_{c,s_p}$ were derived from Bressler et al. (2021) (*11*), which reports national projections across time periods and RCP scenarios referenced to the 2001–2020 average. The year 2010 was used as a reference year to harmonize global and country-level temperature-change baselines across data sources.

**Table S2.** Projected change in global mean surface temperature (°C) for the mid and late 21st century, relative to the 1986-2005 period (provided by IPCC) (*29*).

| Scenario and year | Mean surface temperature (°C) | Likely range (°C) |
|---|---|---|
| RCP 2.6 2040-2059 | 1 | 0.4 to 1.6 |
| RCP 2.6 2080-2099 | 1 | 0.3 to 1.7 |
| RCP 4.5 2040-2059 | 1.4 | 0.9 to 2 |
| RCP 4.5 2080-2099 | 1.8 | 1.1 to 2.6 |
| RCP 6.0 2040-2059 | 1.3 | 0.8 to 1.8 |
| RCP 6.0 2080-2099 | 2.2 | 1.4 to 3.1 |
| RCP 8.5 2040-2059 | 2 | 1.4 to 2.6 |
| RCP 8.5 2080-2099 | 3.7 | 2.6 to 4.8 |

### Characterization factor: Damage per kg of GHGs emitted and Absolute Global Temperature Change Potential

*Characterization factors:* Since the per °C damage factors were largely independent of the RCP scenarios and of the corresponding global average temperature increase (see results figure 2 main text), it is possible to calculate the characterization factors by combining the damage factors of equations S5 and S6 with the latest $AGTP_{cum}$ from AR6. This is in line with the Absolute Global Temperature Change Potential (AGTP) framework introduced by Shine et al. (*30*) and the approach developed by Levasseur et al. (*31*, *32*) to estimate cumulative temperature changes over time following a pulse emission of GHGs, while updating the calculation of $AGTP_{cum}$, accounting for the latest available data (see section 3.1 below).

The characterisation factor for kg of gas $k$ over a specific time horizon h is denoted as $CF_{k,h}$, expressed in [DALY·$kg_{GHG}^{-1}$]. It is calculated as the product of $AGTP_{cum\,k,h}$ expressed in [°C·yr·$kg_{GHG\ k}^{-1}$] and damage factor $DF_p$ expressed in [DALY·°C$^{-1}$·yr$^{-1}$], which represents the country specific damage factor for the chosen time period and is calculated according to Equation S1.

Since aggregated damage factors $DF_{s_p}$ were obtained for two-time horizon periods (0-100 and 100-500 years), we combined them to derive a final characterisation factor $CF_{0-500,k}$ representing the full 0-500 year period, as shown in Equation S7.

$$CF_{c,r,0-500,k} = (DF_{c,r,0-100} \times AGTPcum_{k,0-100}) + DF_{c,r,100-500} \times \left(AGTPcum_{k,0-500} - AGTPcum_{k,0-100}\right)$$

(Equation S7)

The average CFs are given in Table 1 of the main text, and the country specific CFs are detailed in data S4. For the perfluorocarbon group, since emission data are directly given per kgCO2 equivalent, the $CF_{\mathrm{CO2},0\text{–}500}$ is also used for PFCs. Updated CF values used in this study are provided in data S5.

*Determination of $AGTP_{cum}$*

$AGTP_{cum,k}$ is specific to each GHG *k* and time horizon (0-100 or 100-500 years) but is independent of the country, since the country specific DF were calculated per degree of mean global temperature increase (in contrast to a country specific temperature increase). To align our modelling with the latest IPCC assessment, we derived cumulative AGTP for $CO_2$ using the impulse-response formulation of Gasser et al. (*33*) as implemented in the official IPCC WG1 Chapter 7 code. Cumulative AGTP for non-$CO_2$ gases was then inferred from the AR6 GWP values at 100- and 500-year horizons to ensure consistency across gases:

$$AGTPcum_{k,h} = AGTPcum_{CO2,h} \times GWP_{h,k}$$

(Equation S8)

Using the IPCC WG1 GitHub values (https://github.com/IPCC-WG1/Chapter-7) (*34*), our reference $AGTP_{cum}$ ($CO_2$) is $4.35\times10^{-14}$ [°C·yr·kg$^{-1}$] for 0-100 years and $1.95\times10^{-13}$ [°C·yr·kg$^{-1}$] for 0-500 years. Model evaluation shows our AGTP and $AGTP_{cum,k}$ are consistent with the AR6 repository, within 0.3%.

### Damages from present worldwide GHG emissions

Having derived GHG-specific characterization factors $CF_{k,h}$and combined 0-500-year factors $CF_{k,0\text{-}500}$ from aggregated damage factors and cumulative temperature response in Sections 2 and 3, we finally estimate annual health damages attributable to present-day national consumption using **equation 1b**.

The country-specific annual consumption-based emissions $E_{GHG\ k,i}$, obtained from the EORA (*24*), multi-regional input-output database are multiplied by the GHG-specific characterization factors from equation S7 to estimate the induced health damages per country, expressed in $[\text{DALY} \cdot \text{yr}^{-1}]$. Countries whose consumption induces emissions are designated as emission inducers $i$.

*GHG Emission induced per country of consumption*

We compute consumption-based emissions for the six greenhouse gases available in EORA (carbon dioxide **(**$CO_2$), methane fossil ($CH_4$), nitrous oxide ($N_2O$), sulfur hexafluoride ($SF_6$), nitrogen trifluoride (NF3), and perfluorocarbons (PFCs); see Table 1 main text) (*24*). Hydrofluorocarbons were excluded from the consumption-based analysis due to inventory limitations. To quantify annual consumption-based emissions of each greenhouse gas $k$ induced by country $i$, we applied a standard MRIO approach using the EORA database (*24*). These emissions $E_{k,i}$were computed as:

$$E_{GHG\ k,i} = B_{k,j}\ (I - A)^{-1} \times d_i$$

(Equation S9)

where $B_{k,j}$ is the emissions intensity vector for gas $k\ \left[\frac{kg\ GHG}{\text{unit of economic output}}\right] for\ country\ of\ emission\ j$, $(I - A)^{-1}$ is the Leontief inverse capturing total supply-chain requirements associated with final demand, and $d_i$is the final demand vector for country $i$. Here, $I$ is the identity matrix and $A$ is the input–output coefficient matrix (unitless).

While EORA provides multiple satellite accounts for GHG emissions (*24*), we relied on the PRIMAP-hist dataset (*35*). Emissions from land use, land-use change, and forestry (LULUCF) were excluded due to limited data availability and methodological limitations, such that our estimates reflect non-LULUCF anthropogenic emissions.

*Determination of induced health damages in country c for heat and cold attributable to the consumption of country $i$*

GHG-specific induced health damages in country c for heat and cold attributable to the consumption of country $i$ ($D_{r,i \to c}$) are then calculated according to equation by multiplying consumption-based emissions by the corresponding combined characterization factors as described in equation S1. The total damage induced by a country or region is given by the sum of the damage over all receiving countries, as displayed by the stacked bar Figure 3A in the main text. Inversely, the total damage received by a given country or region can be displayed as the sum of the damage created by all inducing regions, as displayed in figure 3B.

**Uncertainty analysis on damage and characterization Factors**

Table S3 summarizes the hypotheses made in the uncertainty analysis. We implemented two computational pathways for the uncertainty propagation, (i) an independent low uncertainty estimate assuming full independency of variables across scenarios and countries, and (ii) a dependent high uncertainty estimate assuming dependency to assess two extreme uncertainty propagation. All simulation used 10,000 Monte Carlo iterations, since additional tests with 100,000 iterations showed negligible differences in summary statistics, confirming that these 10,000 iterations are sufficient.

**Table S3.** Description of the distribution type, determination of the standard errors and of the hypotheses made for the dependent scenario in the uncertainty analysis.

| Variable | Distribution type | Standard error (SE) and source | Hypothesis for dependent scenario |
|---|---|---|---|
| Relative risk<br>RR [%] | Normal | Country specific<br>SE from Bressler 2021 | All country RR samples ranked lowest to highest |
| DALY per death<br>DD [DALY·death$^{-1}$] | Lognormal | GSD=1.095, derived from GBD Death & DALY CIs | Same two DD samples applied across all scenarios |
| Damage factor*<br>DF [DALY·°C$^{-1}$·yr$^{-1}$] | Close to normal distribution | Resulting CIs from Monte-Carlo analysis, final CI taken as average CI ind and dep | All countries, and health risk DF samples ranked to calculate world DFs |
| Cumulative Absolute Global Temperature Change Potential<br>$AGTP_{cum}$ [°C·yr·$kg_{GHG}^{-1}$] | Normal | SE adapted from Ziegler, 2025 (*36*) | $AGTPcum_{k,0-500} - AGTPcum_{k,0-100}$ calculated using the same sampling as $AGTPcum_{k,0-100}$ |
| CF for $CO_2$<br>[DALY· $kg_{CO2}^{-1}$] | Determined by Monte-Carlo | Equation S7, final CF taken as average CF ind and dep | Calculated using ranked samples of AGTPcum & DF |
| GWP100 and GWP500<br>[$kg_{CO2}$· $kg_{GHG}^{-1}$] | Normal distribution | From: IPCC AR5, Chapter 7 Supplementary Material, Tables 7.SM7 to 7.SM13 (*37*) | GWP100 and GWP 500 samples ranked before calculating CFs |
| CF for other GHGs<br>[DALY· $kg_{GHG}^{-1}$] | Determined by Monte-Carlo | Equation S7 and S8 | Using ranked values of CF for CO2 and for GWPs |

*No additional uncertainty was introduced for DTg since uncertainty on temperature increase will be considered in the AGTP, nor for the mortality rate whose uncertainty is limited since it is directly based on observed data.

Uncertainty in the spatially resolved DFs was quantified by propagating variation from two primary sources: (i) country- and scenario-specific temperature-attributable mortality relative risks as parameterized following Bressler et al. (2021) (*11*), and ii) DALY per death (DDc,r). For DALY per death (DDc,r), distributions were derived comparing GBD uncertainties between death and DALYs; consistent with the small average increase in uncertainty when moving from deaths to DALYs for non-optimal temperature (≈ 2% on average across heat and cold), we modeled DDc,r as a lognormal distribution, with a geometric standard deviation (GSD) of 1.095. In the independent pathway, heat- and cold-specific DDc,r values were sampled independently for each of the eight scenario combinations defined in the study within a given Monte Carlo iteration. In the dependent uncertainty pathway, the same two sampled DDc,r values (one for heat, one for cold) were applied across all applicable scenario combinations within that iteration, yielding a more conservative spread that avoids compounding scenario-level sampling. For the mortality-change parameters, the independent uncertainties were sampled independently for each country and scenario specific values (heat, cold, and heat with adaptation) from normal distributions, independently across countries and scenarios, forming the baseline ("independent") uncertainty structure.

To represent a plausible extreme of shared model bias across countries and scenarios, we conducted the additional dependent run by first sorting, for each country and scenario, their mortality rate (i.e. the lowest value for iteration 1, the second lowest for iteration 2, etc.) This introduces extreme iterations where each country and scenario yields their best and worst outcome. Consistency checks verified that DF probabilistic medians are consistent with deterministic DF values, confirming correct propagation and export of sampled parameters (within 10%, Table S1). Uncertainty intervals for damage factors can be found in data S4.

This ranking approach was then repeated as described in the last column of Table S3 for determining the AGTPcum and the characterization factors. For AGTP cum, we assumed normal distribution with a standard deviation derived from Zieger et al. (2025, table 3) (*36*). Uncertainties on the Global Warming Potential 100 and 500 years were determined assuming normal distribution and standard error derived from the IPCC WG1 GitHub values (https://github.com/IPCC-WG1/Chapter-7). For GHGs not explicitly mentioned in this table, standard errors of 38% for GP100 and 40% for GWP500 were retained as the average value for all GHGs but $CH_4$ and $N_2O$. The final uncertainties of the damage and characterization factors were calculated as the average between the low end independent and the high-end dependent 95$^{th}$ confidence limits and are presented in Table S1 above and in Table 1 of the main text.

The country specific net adjusted damage factors with both dependent and independent confidence interval are presented in figure S2 for shorter term 0-100 years and in figure S3 for long-term 100-500 years, ranked from lowest (Kenya) to highest DF (Niger). As expected, the dependent uncertainties are larger than the independent calculation by up to a factor 2. Despite these high dependent uncertainties, the calculated country specific damage factors are nevertheless significantly higher than zero for all countries with most substantial impacts. The average between dependent and independent was used for the calculations of final uncertainties (figures S4 and S5).Very similar patterns are observed at the level of characterization factors for the average net (figure S6) and net adjusted characterization factors per capita receiving countries (figure S7), with the highest CFs in Sierra Leone, Mali, Chad, Central African Republic, and Niger (countries 159 to 163). China (87) and India (130) have substantially lower CFs per capita. When multiplying by the population of each country, the cumulative impacts due to heat and cold per kg CO2 emitted is the highest in India due to its large population and moderate CF. Inversely, the cumulative impact for Central African Republic who was the second highest per capita is only ranked 130 out of the 163 countries. Niger, which had the highest impact per capita with a still important population of 23 million, is now ranked 5$^{th}$ highest country for the cumulative impacts. Comprehensive uncertainty results and country specific graphs can be found in the different sheets of data S4.

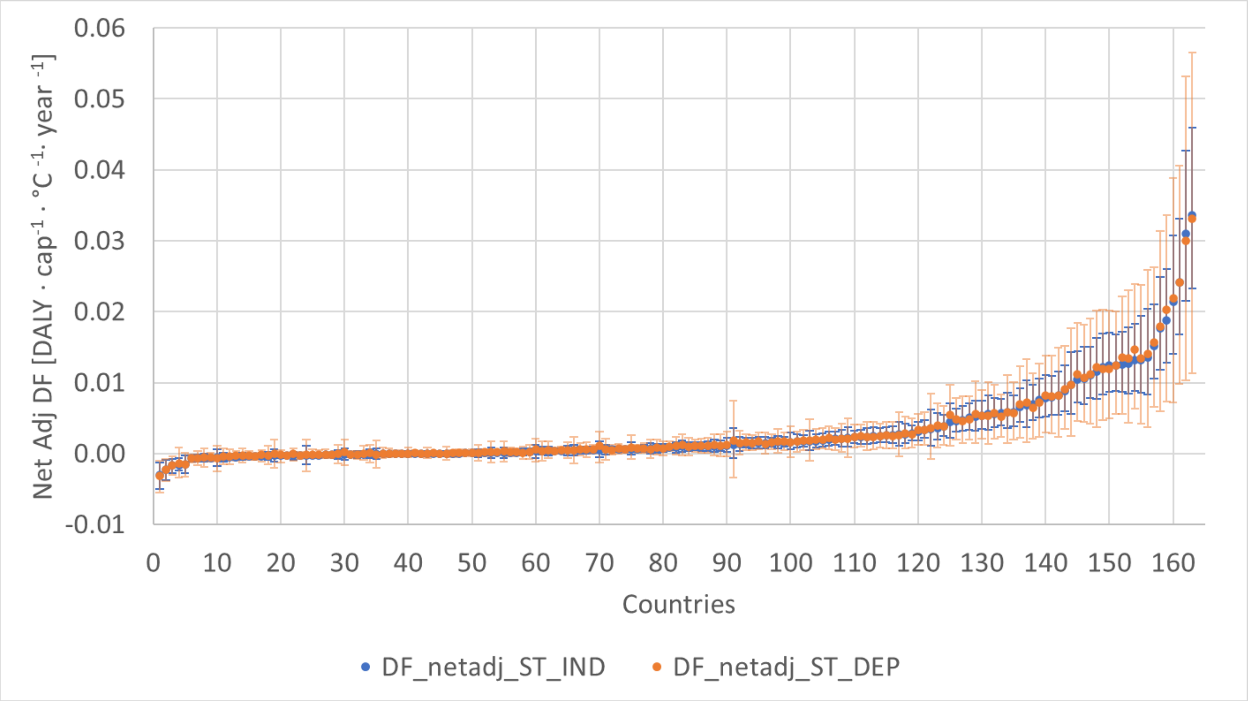


**Fig. S2.** Net adjusted, independent (blue) and dependent (orange), shorter-term 0-100 years damage factors for each country, with countries sorted from lowest (Kenya) to highest (Niger) value.

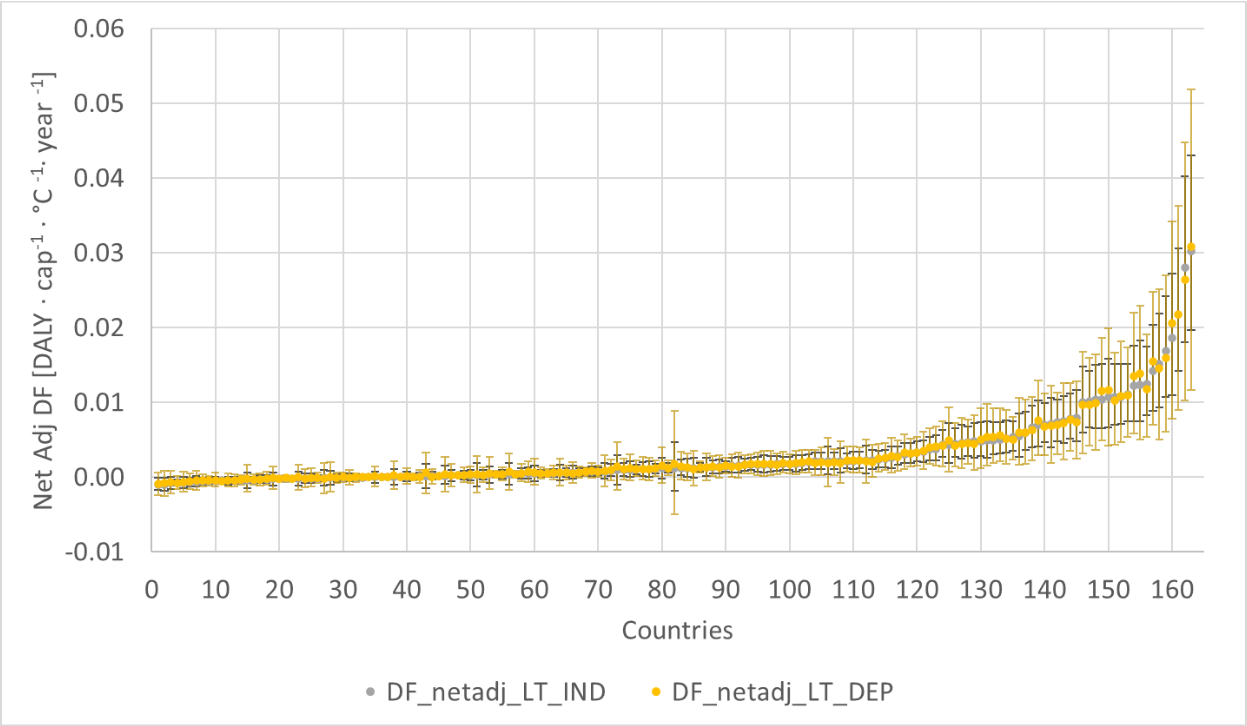


**Fig. S3.** Net adjusted, independent (brown) and dependent (yellow), long-term 100-500 years, damage factors per capita receiving countries, with countries sorted from lowest (Kenya) to highest (Niger) value.

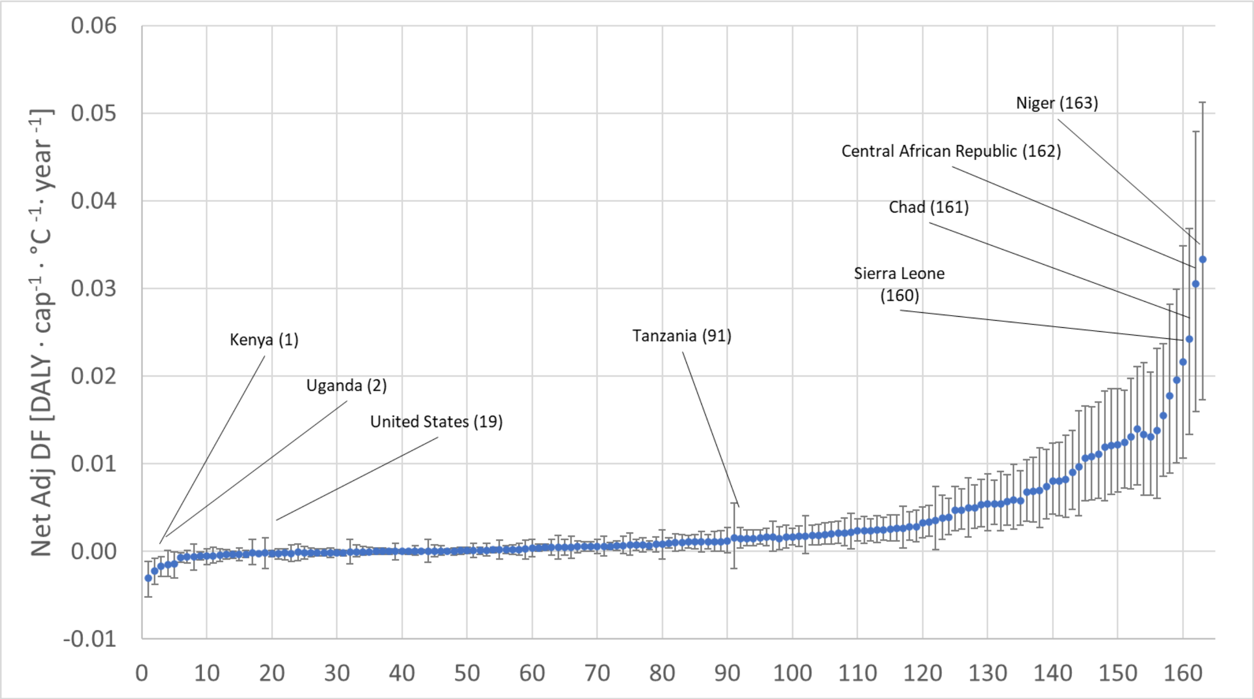


**Fig. S4.** Net adjusted shorter term 0-100 years, damage factors per capita receiving countries, for the average between dependent and independent uncertainties, sorted from lowest (Kenya) to highest (Niger) value.

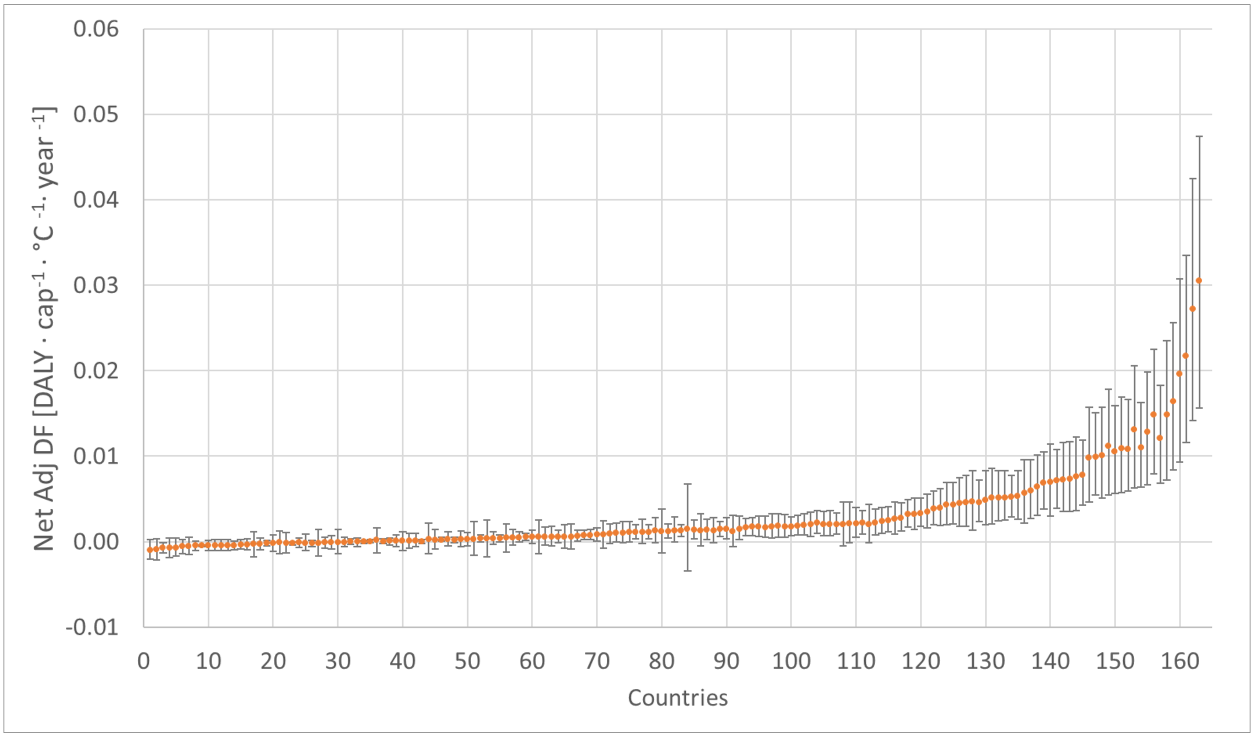


**Fig. S5.** Net adjusted long-term 100-500 years, damage factors per capita receiving countries, for the average between dependent and independent uncertainties, from lowest (Sweden) to highest (Niger) value.

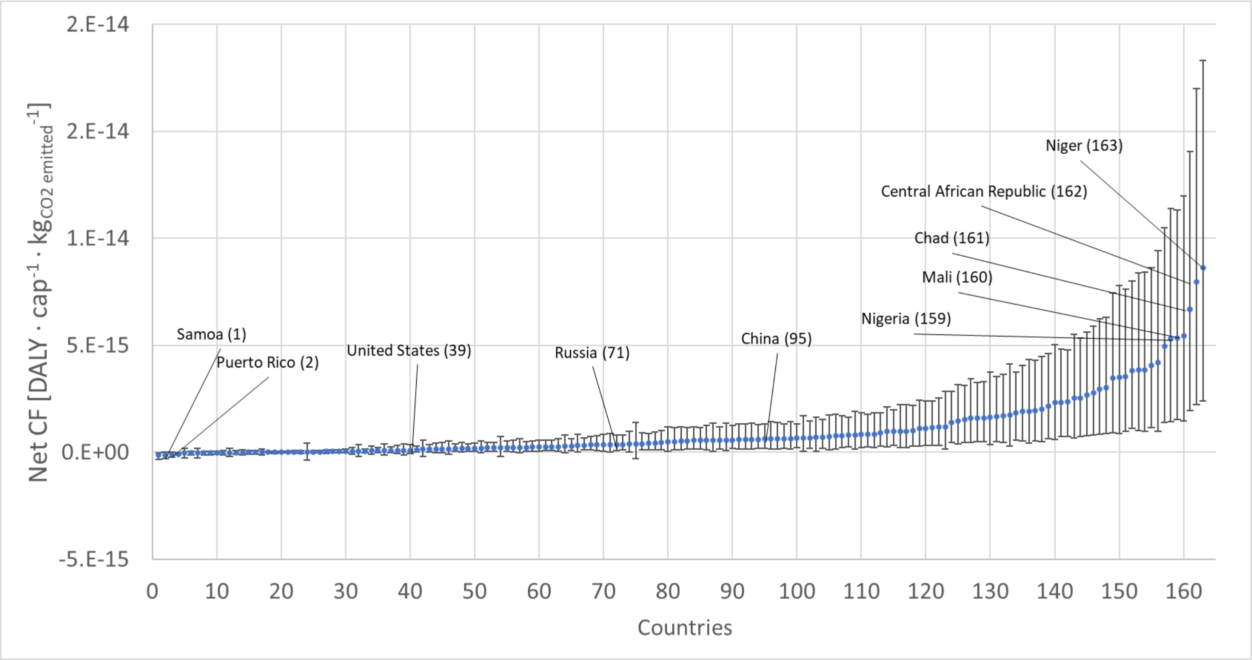


**Fig. S6.** Net average characterization factors per capita receiving countries for $CO_2$, from lowest (Sweden) to highest (Niger) value.

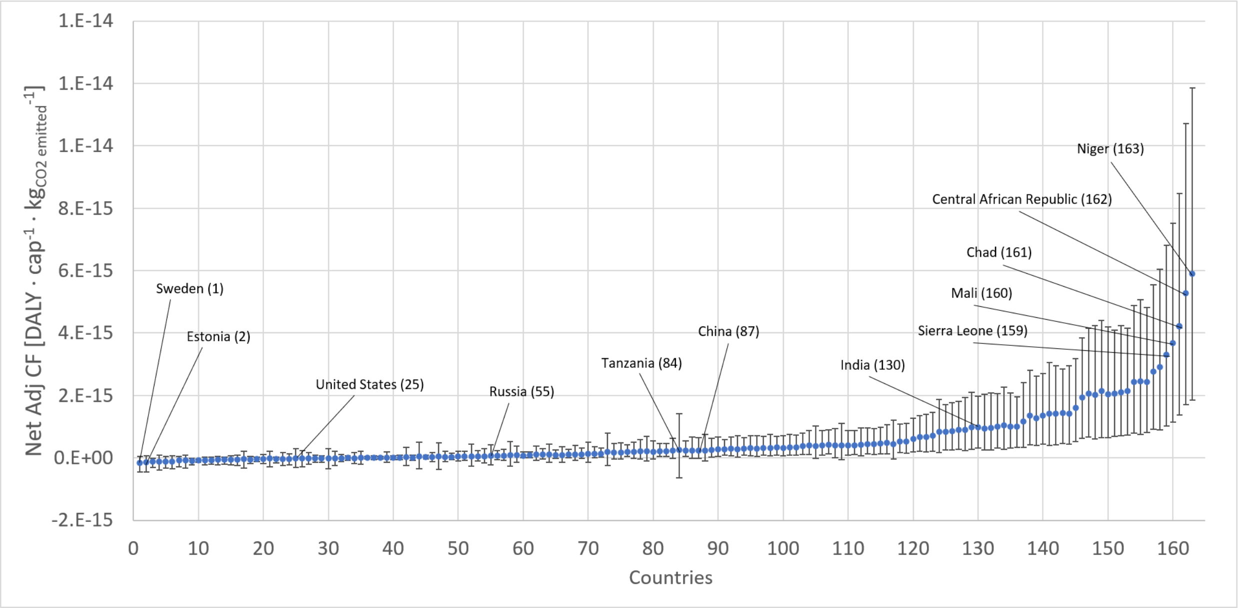


**Fig. S7.** Net adjusted average characterization factors per capita receiving countries for $CO_2$, from lowest (Samoa) to highest (Niger) value.

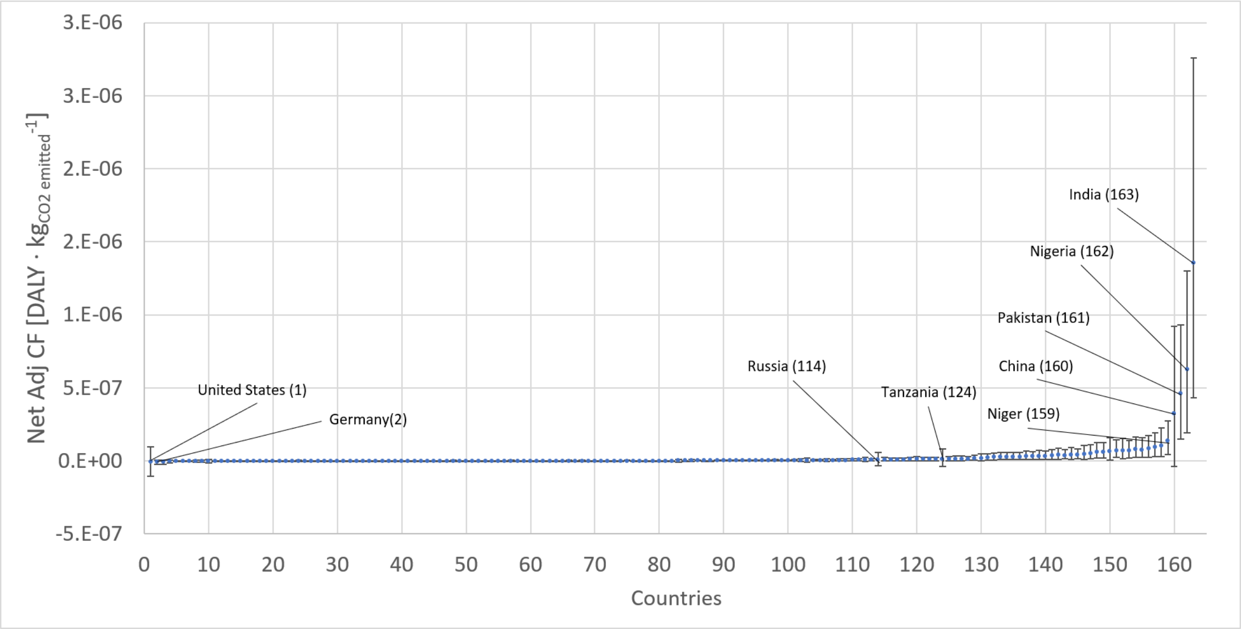


**Fig. S8.** Net adjusted, average cumulative characterization factors per receiving countries for $CO_2$, from lowest (USA) to highest (India) value.

4. M. Romanello, M. Walawender, S.-C. Hsu, A. Moskeland, Y. Palmeiro-Silva, D. Scamman, Z. Ali, N. Ameli, D. Angelova, S. Ayeb-Karlsson, S. Basart, J. Beagley, P. J. Beggs, L. Blanco-Villafuerte, W. Cai, M. Callaghan, D. Campbell-Lendrum, J. D. Chambers, V. Chicmana-Zapata, L. Chu, T. J. Cross, K. R. van Daalen, C. Dalin, N. Dasandi, S. Dasgupta, M. Davies, R. Dubrow, M. J. Eckelman, J. D. Ford, C. Freyberg, O. Gasparyan, G. Gordon-Strachan, M. Grubb, S. H. Gunther, I. Hamilton, Y. Hang, R. Hänninen, S. Hartinger, K. He, J. Heidecke, J. J. Hess, L. Jamart, S. Jankin, H. Jatkar, O. Jay, I. Kelman, H. Kennard, G. Kiesewetter, P. Kinney, D. Kniveton, R. Kouznetsov, P. Lampard, J. K. W. Lee, B. Lemke, B. Li, Y. Liu, Z. Liu, A. Llabrés-Brustenga, M. Lott, R. Lowe, J. Martinez-Urtaza, M. Maslin, L. McAllister, C. McMichael, Z. Mi, J. Milner, K. Minor, J. Minx, N. Mohajeri, N. C. Momen, M. Moradi-Lakeh, K. Morrisey, S. Munzert, K. A. Murray, N. Obradovich, M. B. O'Hare, C. Oliveira, T. Oreszczyn, M. Otto, F. Owfi, O. L. Pearman, F. Pega, A. J. Perishing, A.-C. Pinho-Gomes, J. Ponmattam, M. Rabbaniha, J. Rickman, E. Robinson, J. Rocklöv, D. Rojas-Rueda, R. N. Salas, J. C. Semenza, J. D. Sherman, J. Shumake-Guillemot, P. Singh, H. Sjödin, J. Slater, M. Sofiev, C. Sorensen, M. Springmann, Z. Stalhandske, J. D. Stowell, M. Tabatabaei, J. Taylor, D. Tong, C. Tonne, M. Treskova, J. A. Trinanes, A. Uppstu, F. Wagner, L. Warnecke, H. Whitcombe, P. Xian, C. Zavaleta-Cortijo, C. Zhang, R. Zhang, S. Zhang, Y. Zhang, Q. Zhu, P. Gong, H. Montgomery, A. Costello, The 2024 report of the Lancet Countdown on health and climate change: facing record-breaking threats from delayed action. *The Lancet* **404**, 1847–1896 (2024).

5. K. R. Smith, A. Woodward, D. Campbell-Lendrum, D. D. Chadee, Y. Honda, Q. Liu, J. M. Olwoch, B. Revich, R. Sauerborn, "Human health: impacts, adaptation, and co-benefits" in *Climate Change 2014: Impacts, Adaptation, and Vulnerability. Part A: Global and Sectoral Aspects. Contribution of Working Group II to the Fifth Assessment Report of the Intergovernmental Panel on Climate Change*, C. B. Field, V. R. Barros, D. J. Dokken, K. J. Mach, M. D. Mastrandrea, T. E. Bilir, M. Chatterjee, K. L. Ebi, Y. O. Estrada, R. C.

Genova, B. Girma, E. S. Kissel, A. N. Levy, S. MacCracken, P. R. Mastrandrea, L. L. White, Eds. (Cambridge University Press, Cambridge, UK and New York, NY, USA, 2014), pp. 709–754.

6. C. J. L. Murray, A. D. Lopez, Eds., *The Global Burden of Disease: A Comprehensive Assessment of Mortality and Disability from Diseases, Injuries, and Risk Factors in 1990 and Projected to 2020* (Harvard School of Public Health, Boston, 1996).

7. G. Bianco, R. M. Espinoza-Chávez, P. G. Ashigbie, H. Junio, C. Borhani, S. Miles-Richardson, J. Spector, Projected impact of climate change on human health in low- and middle-income countries: a systematic review. *BMJ Glob Health* **8** (2024).

8. S. Schöngart, Z. Nicholls, R. Hoffmann, S. Pelz, C.-F. Schleussner, High-income groups disproportionately contribute to climate extremes worldwide. *Nat. Clim. Chang.* **15**, 627–633 (2025).

9. A. Gasparrini, Y. Guo, F. Sera, A. M. Vicedo-Cabrera, V. Huber, S. Tong, M. de S. Z. S. Coelho, P. H. N. Saldiva, E. Lavigne, P. M. Correa, N. V. Ortega, H. Kan, S. Osorio, J. Kyselý, A. Urban, J. J. K. Jaakkola, N. R. I. Ryti, M. Pascal, P. G. Goodman, A. Zeka, P. Michelozzi, M. Scortichini, M. Hashizume, Y. Honda, M. Hurtado-Diaz, J. C. Cruz, X. Seposo, H. Kim, A. Tobias, C. Iñiguez, B. Forsberg, D. O. Åström, M. S. Ragettli, Y. L. Guo, C. Wu, A. Zanobetti, J. Schwartz, M. L. Bell, T. N. Dang, D. D. Van, C. Heaviside, S. Vardoulakis, S. Hajat, A. Haines, B. Armstrong, Projections of temperature-related excess mortality under climate change scenarios. *The Lancet Planetary Health* **1**, e360–e367 (2017).

10. Y. Guo, A. Gasparrini, S. Li, F. Sera, A. M. Vicedo-Cabrera, M. de S. Z. S. Coelho, P. H. N. Saldiva, E. Lavigne, B. Tawatsupa, K. Punnasiri, A. Overcenco, P. M. Correa, N. V. Ortega, H. Kan, S. Osorio, J. J. K. Jaakkola, N. R. I. Ryti, P. G. Goodman, A. Zeka, P. Michelozzi, M. Scortichini, M. Hashizume, Y. Honda, X. Seposo, H. Kim, A. Tobias, C. Íñiguez, B. Forsberg, D. O. Åström, Y. L. Guo, B.-Y. Chen, A. Zanobetti, J. Schwartz, T. N. Dang, D. D. Van, M. L. Bell, B. Armstrong, K. L. Ebi, S. Tong, Quantifying excess deaths related to heatwaves under climate change scenarios: A multicountry time series modelling study. *PLOS Medicine* **15**, e1002629 (2018).

11. R. D. Bressler, F. C. Moore, K. Rennert, D. Anthoff, Estimates of country level temperature-related mortality damage functions. *Sci Rep* **11**, 20282 (2021).

12. Y. Honda, M. Kondo, G. McGregor, H. Kim, Y.-L. Guo, Y. Hijioka, M. Yoshikawa, K. Oka, S. Takano, S. Hales, R. S. Kovats, Heat-related mortality risk model for climate change impact projection. *Environ Health Prev Med* **19**, 56–63 (2014).

13. A. M. Vicedo-Cabrera, N. Scovronick, F. Sera, D. Royé, R. Schneider, A. Tobias, C. Astrom, Y. Guo, Y. Honda, D. M. Hondula, R. Abrutzky, S. Tong, M. de S. Z. S. Coelho, P. H. N. Saldiva, E. Lavigne, P. M. Correa, N. V. Ortega, H. Kan, S. Osorio, J. Kyselý, A. Urban, H. Orru, E. Indermitte, J. J. K. Jaakkola, N. Ryti, M. Pascal, A. Schneider, K. Katsouyanni, E. Samoli, F. Mayvaneh, A. Entezari, P. Goodman, A. Zeka, P. Michelozzi,

F. de'Donato, M. Hashizume, B. Alahmad, M. H. Diaz, C. D. L. C. Valencia, A. Overcenco, D. Houthuijs, C. Ameling, S. Rao, F. Di Ruscio, G. Carrasco-Escobar, X. Seposo, S. Silva, J. Madureira, I. H. Holobaca, S. Fratianni, F. Acquaotta, H. Kim, W. Lee, C. Iniguez, B. Forsberg, M. S. Ragettli, Y. L. L. Guo, B. Y. Chen, S. Li, B. Armstrong, A. Aleman, A. Zanobetti, J. Schwartz, T. N. Dang, D. V. Dung, N. Gillett, A. Haines, M. Mengel, V. Huber, A. Gasparrini, The burden of heat-related mortality attributable to recent human-induced climate change. *Nat. Clim. Chang.* **11**, 492–500 (2021).

14. M. Ezzati, A. D. Lopez, A. Rodgers, C. J. L. Murray, Eds., *Comparative Quantification of Health Risks: Global and Regional Burden of Disease Attributable to Selected Major Risk Factors* (World Health Organization, Geneva, 2004).

21. K. Riahi, D. P. van Vuuren, E. Kriegler, J. Edmonds, B. C. O'Neill, S. Fujimori, N. Bauer, K. Calvin, R. Dellink, O. Fricko, W. Lutz, A. Popp, J. C. Cuaresma, S. Kc, M. Leimbach, L. Jiang, T. Kram, S. Rao, J. Emmerling, K. Ebi, T. Hasegawa, P. Havlik, F. Humpenöder, L. A. Da Silva, S. Smith, E. Stehfest, V. Bosetti, J. Eom, D. Gernaat, T. Masui, J. Rogelj, J. Strefler, L. Drouet, V. Krey, G. Luderer, M. Harmsen, K. Takahashi, L. Baumstark, J. C. Doelman, M. Kainuma, Z. Klimont, G. Marangoni, H. Lotze-Campen, M. Obersteiner, A. Tabeau, M. Tavoni, The Shared Socioeconomic Pathways and their energy, land use, and greenhouse gas emissions implications: An overview. *Global Environmental Change* **42**, 153–168 (2017).

22. T. Vos, S. S. Lim, C. Abbafati, K. M. Abbas, M. Abbasi, M. Abbasifard, M. Abbasi-Kangevari, H. Abbastabar, F. Abd-Allah, A. Abdelalim, M. Abdollahi, I. Abdollahpour, H. Abolhassani, V. Aboyans, E. M. Abrams, L. G. Abreu, M. R. M. Abrigo, L. J. Abu-Raddad, A. I. Abushouk, A. Acebedo, I. N. Ackerman, M. Adabi, A. A. Adamu, O. M. Adebayo, V. Adekanmbi, J. D. Adelson, O. O. Adetokunboh, D. Adham, M. Afshari, A. Afshin, E. E. Agardh, G. Agarwal, K. M. Agesa, M. Aghaali, S. M. K. Aghamir, A. Agrawal, T. Ahmad, A. Ahmadi, M. Ahmadi, H. Ahmadieh, E. Ahmadpour, T. Y. Akalu, R. O. Akinyemi, T. Akinyemiju, B. Akombi, Z. Al-Aly, K. Alam, N. Alam, S. Alam, T. Alam, T. M. Alanzi, S. B. Albertson, J. E. Alcalde-Rabanal, N. M. Alema, M. Ali, S. Ali, G. Alicandro, M. Alijanzadeh, C. Alinia, V. Alipour, S. M. Aljunid, F. Alla, P. Allebeck, A. Almasi-Hashiani, J. Alonso, R. M. Al-Raddadi, K. A. Altirkawi, N. Alvis-Guzman, N. J. Alvis-Zakzuk, S. Amini, M. Amini-Rarani, A. Aminorroaya, F. Amiri, A. M. L. Amit, D. A. Amugsi, G. G. H. Amul, D. Anderlini, C. L. Andrei, T. Andrei, M. Anjomshoa, F. Ansari, I. Ansari, A. Ansari-Moghaddam, C. A. T. Antonio, C. M. Antony, E. Antriyandarti, D. Anvari, R. Anwer, J. Arabloo, M. Arab-Zozani, A. Y. Aravkin, F. Ariani, J. Ärnlöv, K. K. Aryal, A. Arzani, M. Asadi-Aliabadi, A. A. Asadi-Pooya, B. Asghari, C. Ashbaugh, D. D. Atnafu, S. R. Atre, F. Ausloos, M. Ausloos, B. P. A. Quintanilla, G. Ayano, M. A. Ayanore, Y. A. Aynalem, S. Azari, G. Azarian, Z. N. Azene, E. Babaee, A. Badawi, M. Bagherzadeh, M. H. Bakhshaei, A. Bakhtiari, S. Balakrishnan, S. Balalla, S. Balassyano, M. Banach, P. C. Banik, M. S. Bannick, A. B. Bante, A. G. Baraki, M. A. Barboza, S. L. Barker-Collo, C. M. Barthelemy, L. Barua, A. Barzegar, S. Basu, B. T. Baune, M. Bayati, G. Bazmandegan, N. Bedi, E. Beghi, Y. Béjot, A. K. Bello, R. G. Bender, D. A. Bennett, F. B. Bennitt, I. M. Bensenor, C. P. Benziger, K. Berhe, E. Bernabe, G. J. Bertolacci, R. Bhageerathy, N. Bhala, D. Bhandari, P. Bhardwaj, K. Bhattacharyya, Z. A. Bhutta, S. Bibi, M. H. Biehl, B. Bikbov, M. S. B. Sayeed, A. Biondi, B. M. Birihane, D. Bisanzio, C. Bisignano, R. K. Biswas, S. Bohlouli, M. Bohluli, S. R. R. Bolla, A. Boloor, A. S. Boon-Dooley, G. Borges, A. M. Borzì, R. Bourne, O. J. Brady, M. Brauer, C. Brayne, N. J. K. Breitborde, H. Brenner, P. S. Briant, A. M. Briggs, N. I. Briko, G. B. Britton, D. Bryazka, R. Buchbinder, B. R. Bumgarner, R. Busse, Z. A. Butt, F. L. C. dos Santos, L. L. A. Cámera, I. R. Campos-Nonato, J. Car, R. Cárdenas, G. Carreras, J. J. Carrero, F. Carvalho, J. M. Castaldelli-Maia, C. A. Castañeda-Orjuela, G. Castelpietra, C. D. Castle, F. Castro, F. Catalá-López, K. Causey, C. R. Cederroth, K. M. Cercy, E. Cerin, J. S. Chandan, A. R. Chang, F. J. Charlson, V. K. Chattu, S. Chaturvedi, O. Chimed-Ochir, K. L. Chin, D. Y. Cho, H. Christensen, D.-T. Chu, M. T. Chung, F. M. Cicuttini, L. G. Ciobanu, M. Cirillo, E. L. Collins, K. Compton, S. Conti, P. A. Cortesi, V. M. Costa, E. Cousin, R. G. Cowden, B. C. Cowie, E. A. Cromwell, D. H. Cross, C. S. Crowe, J. A. Cruz, M. Cunningham, S. M. A. Dahlawi, G. Damiani, L. Dandona, R. Dandona, A. M. Darwesh, A. Daryani, J. K. Das, R. D. Gupta, J. das Neves, C. A. Dávila-Cervantes, K. Davletov, D. D. Leo, F. E. Dean, N. K. DeCleene, A. Deen, L. Degenhardt, R. P. Dellavalle, F. M. Demeke, D. G. Demsie, E. Denova-Gutiérrez, N. D. Dereje, N. Dervenis, R. Desai, A. Desalew, G. A. Dessie, S. D. Dharmaratne, G. P. Dhungana, M. Dianatinasab, D. Diaz, Z. S. D. Forooshani, Z. V. Dingels, M. A. Dirac, S. Djalalinia, H. T. Do, K. Dokova, F. Dorostkar, C. P. Doshi, L. Doshmangir, A. Douiri, M. C. Doxey, T. R. Driscoll, S. J. Dunachie, B. B. Duncan, A. R. Duraes, A. W. Eagan, M. E. Kalan, D. Edvardsson, J. R. Ehrlich, N. E. Nahas, I. E. Sayed, M. E. Tantawi, I. Elbarazi, I. Y. Elgendy, H. R. Elhabashy, S. I. El-Jaafary, I. R. Elyazar, M. H. Emamian, S. Emmons-Bell, H. E. Erskine, B. Eshrati, S. Eskandarieh, S.

Esmaeilnejad, F. Esmaeilzadeh, A. Esteghamati, K. Estep, A. Etemadi, A. E. Etisso, M. Farahmand, A. Faraj, M. Fareed, R. Faridnia, C. S. e S. Farinha, A. Farioli, A. Faro, M. Faruque, F. Farzadfar, N. Fattahi, M. Fazlzadeh, V. L. Feigin, R. Feldman, S.-M. Fereshtehnejad, E. Fernandes, A. J. Ferrari, M. L. Ferreira, I. Filip, F. Fischer, J. L. Fisher, R. Fitzgerald, C. Flohr, L. S. Flor, N. A. Foigt, M. O. Folayan, L. M. Force, C. Fornari, M. Foroutan, J. T. Fox, M. Freitas, W. Fu, T. Fukumoto, J. M. Furtado, M. M. Gad, E. Gakidou, N. C. Galles, S. Gallus, A. Gamkrelidze, A. L. Garcia-Basteiro, W. M. Gardner, B. S. Geberemariyam, A. M. Gebrehiwot, K. B. Gebremedhin, A. A. A. A. Gebreslassie, A. G. Hayoon, P. W. Gething, M. Ghadimi, K. Ghadiri, M. Ghafourifard, A. Ghajar, F. Ghamari, A. Ghashghaee, H. Ghiasvand, N. Ghith, A. Gholamian, S. A. Gilani, P. S. Gill, M. Gitimoghaddam, G. Giussani, S. Goli, R. S. Gomez, S. V. Gopalani, G. Gorini, T. M. Gorman, H. C. Gottlich, H. Goudarzi, A. C. Goulart, B. N. G. Goulart, A. Grada, M. Grivna, G. Grosso, M. I. M. Gubari, H. C. Gugnani, A. L. S. Guimaraes, R. A. Guimarães, R. A. Guled, G. Guo, Y. Guo, R. Gupta, J. A. Haagsma, B. Haddock, N. Hafezi-Nejad, A. Hafiz, H. Hagins, L. M. Haile, B. J. Hall, I. Halvaei, R. R. Hamadeh, K. H. Abdullah, E. B. Hamilton, C. Han, H. Han, G. J. Hankey, J. M. Haro, J. D. Harvey, A. I. Hasaballah, A. Hasanzadeh, M. Hashemian, S. Hassanipour, H. Hassankhani, R. J. Havmoeller, R. J. Hay, S. I. Hay, K. Hayat, B. Heidari, G. Heidari, R. Heidari-Soureshjani, D. Hendrie, H. J. Henrikson, N. J. Henry, C. Herteliu, F. Heydarpour, T. R. Hird, H. W. Hoek, M. K. Hole, R. Holla, P. Hoogar, H. D. Hosgood, M. Hosseinzadeh, M. Hostiuc, S. Hostiuc, M. Househ, D. G. Hoy, M. Hsairi, V. C. Hsieh, G. Hu, T. M. Huda, F. N. Hugo, C. K. Huynh, B.-F. Hwang, V. C. Iannucci, S. E. Ibitoye, K. S. Ikuta, O. S. Ilesanmi, I. M. Ilic, M. D. Ilic, L. R. Inbaraj, H. Ippolito, S. S. N. Irvani, M. M. Islam, M. Islam, S. M. S. Islam, F. Islami, H. Iso, R. Q. Ivers, C. C. D. Iwu, I. O. Iyamu, J. Jaafari, K. H. Jacobsen, F. Jadidi-Niaragh, H. Jafari, M. Jafarinia, D. Jahagirdar, M. A. Jahani, N. Jahanmehr, M. Jakovljevic, A. Jalali, F. Jalilian, S. L. James, H. Janjani, M. D. Janodia, A. U. Jayatilleke, P. Jeemon, E. Jenabi, R. P. Jha, V. Jha, J. S. Ji, P. Jia, O. John, Y. O. John-Akinola, C. O. Johnson, S. C. Johnson, J. B. Jonas, T. Joo, A. Joshi, J. J. Jozwiak, M. Jürisson, A. Kabir, Z. Kabir, H. Kalani, R. Kalani, L. R. Kalankesh, R. Kalhor, Z. Kamiab, T. Kanchan, B. K. Matin, A. Karch, M. A. Karim, S. E. Karimi, G. M. Kassa, N. J. Kassebaum, S. V. Katikireddi, N. Kawakami, G. A. Kayode, S. H. Keddie, C. Keller, M. Kereselidze, M. A. Khafaie, N. Khalid, M. Khan, K. Khatab, M. M. Khater, M. N. Khatib, M. Khayamzadeh, M. T. Khodayari, R. Khundkar, N. Kianipour, C. Kieling, D. Kim, Y.-E. Kim, Y. J. Kim, R. W. Kimokoti, A. Kisa, S. Kisa, K. Kissimova-Skarbek, M. Kivimäki, C. J. Kneib, A. K. S. Knudsen, J. M. Kocarnik, T. Kolola, J. A. Kopec, S. Kosen, P. A. Koul, A. Koyanagi, M. A. Kravchenko, K. Krishan, K. J. Krohn, B. K. Defo, B. K. Bicer, G. A. Kumar, M. Kumar, P. Kumar, V. Kumar, G. Kumaresh, O. P. Kurmi, D. Kusuma, H. H. Kyu, C. L. Vecchia, B. Lacey, D. K. Lal, R. Lalloo, J. O. Lam, F. H. Lami, I. Landires, J. J. Lang, V. C. Lansingh, S. L. Larson, A. O. Larsson, S. Lasrado, Z. S. Lassi, K. M.-M. Lau, P. M. Lavados, J. V. Lazarus, J. R. Ledesma, P. H. Lee, S. W. H. Lee, K. E. LeGrand, J. Leigh, M. Leonardi, H. Lescinsky, J. Leung, M. Levi, S. Lewington, S. Li, L.-L. Lim, C. Lin, R.-T. Lin, C. Linehan, S. Linn, H.-C. Liu, S. Liu, Z. Liu, K. J. Looker, A. D. Lopez, P. D. Lopukhov, S. Lorkowski, P. A. Lotufo, T. C. D. Lucas, A. Lugo, R. Lunevicius, R. A. Lyons, J. Ma, J. H. MacLachlan, E. R. Maddison, R. Maddison, F. Madotto, P. W. Mahasha, H. T. Mai, A. Majeed, V. Maled, S. Maleki, R. Malekzadeh, D. C. Malta, A. A. Mamun, A. Manafi, N. Manafi, H. Manguerra, B. Mansouri, M. A. Mansournia, A. M. M. Herrera, J. C. Maravilla, A. Marks,

F. R. Martins-Melo, I. Martopullo, S. Z. Masoumi, J. Massano, B. B. Massenburg, M. R. Mathur, P. K. Maulik, C. McAlinden, J. J. McGrath, M. McKee, M. M. Mehndiratta, F. Mehri, K. M. Mehta, W. B. Meitei, P. T. N. Memiah, W. Mendoza, R. G. Menezes, E. W. Mengesha, M. B. Mengesha, A. Mereke, A. Meretoja, T. J. Meretoja, T. Mestrovic, B. Miazgowski, T. Miazgowski, I. M. Michalek, K. M. Mihretie, T. R. Miller, E. J. Mills, A. Mirica, E. M. Mirrakhimov, H. Mirzaei, M. Mirzaei, M. Mirzaei-Alavijeh, A. T. Misganaw, P. Mithra, B. Moazen, M. Moghadaszadeh, E. Mohamadi, D. K. Mohammad, Y. Mohammad, N. M. G. Mezerji, A. Mohammadian-Hafshejani, N. Mohammadifard, R. Mohammadpourhodki, S. Mohammed, A. H. Mokdad, M. Molokhia, N. C. Momen, L. Monasta, S. Mondello, M. D. Mooney, M. Moosazadeh, G. Moradi, M. Moradi, M. Moradi-Lakeh, R. Moradzadeh, P. Moraga, L. Morales, L. Morawska, I. M. Velásquez, J. Morgado-da-Costa, S. D. Morrison, J. F. Mosser, S. Mouodi, S. M. Mousavi, A. M. Khaneghah, U. O. Mueller, S. B. Munro, M. K. Muriithi, K. I. Musa, S. Muthupandian, M. Naderi, A. J. Nagarajan, G. Nagel, B. Naghshtabrizi, S. Nair, A. K. Nandi, V. Nangia, J. R. Nansseu, V. C. Nayak, J. Nazari, I. Negoi, R. I. Negoi, H. B. N. Netsere, J. W. Ngunjiri, C. T. Nguyen, J. Nguyen, M. Nguyen, M. Nguyen, E. Nichols, D. Nigatu, Y. T. Nigatu, R. Nikbakhsh, M. R. Nixon, C. A. Nnaji, S. Nomura, B. Norrving, J. J. Noubiap, C. Nowak, V. Nunez-Samudio, A. Oţoiu, B. Oancea, C. M. Odell, F. A. Ogbo, I.-H. Oh, E. W. Okunga, M. Oladnabi, A. T. Olagunju, B. O. Olusanya, J. O. Olusanya, M. M. Oluwasanu, A. O. Bali, M. O. Omer, K. L. Ong, O. E. Onwujekwe, A. U. Orji, H. M. Orpana, A. Ortiz, S. M. Ostroff, N. Otstavnov, S. S. Otstavnov, S. Øverland, M. O. Owolabi, M. P. A, J. R. Padubidri, A. P. Pakhare, R. Palladino, A. Pana, S. Panda-Jonas, A. Pandey, E.-K. Park, P. G. K. Parmar, D. K. Pasupula, S. K. Patel, A. J. Paternina-Caicedo, A. Pathak, M. Pathak, S. B. Patten, G. C. Patton, D. Paudel, H. P. Toroudi, A. E. Peden, A. Pennini, V. C. F. Pepito, E. K. Peprah, A. Pereira, D. M. Pereira, N. Perico, H. Q. Pham, M. R. Phillips, D. M. Pigott, T. Pilgrim, T. M. Pilz, M. Pirsaheb, O. Plana-Ripoll, D. Plass, K. N. Pokhrel, R. V. Polibin, S. Polinder, K. R. Polkinghorne, M. J. Postma, H. Pourjafar, F. Pourmalek, R. P. Kalhori, A. Pourshams, A. Poznańska, S. I. Prada, V. Prakash, D. R. A. Pribadi, E. Pupillo, Z. Q. Syed, M. Rabiee, N. Rabiee, A. Radfar, A. Rafiee, A. Rafiei, A. Raggi, A. Rahimi-Movaghar, M. A. Rahman, A. Rajabpour-Sanati, F. Rajati, K. Ramezanzadeh, C. L. Ranabhat, P. C. Rao, S. J. Rao, D. Rasella, P. Rastogi, P. Rathi, D. L. Rawaf, S. Rawaf, L. Rawal, C. Razo, S. B. Redford, R. C. Reiner, N. Reinig, M. B. Reitsma, G. Remuzzi, V. Renjith, A. M. N. Renzaho, S. Resnikoff, N. Rezaei, M. sadegh Rezai, A. Rezapour, P.-A. Rhinehart, S. M. Riahi, A. L. P. Ribeiro, D. C. Ribeiro, D. Ribeiro, J. Rickard, N. L. S. Roberts, S. Roberts, S. R. Robinson, L. Roever, S. Rolfe, L. Ronfani, G. Roshandel, G. A. Roth, E. Rubagotti, S. F. Rumisha, S. Sabour, P. S. Sachdev, B. Saddik, E. Sadeghi, M. Sadeghi, S. Saeidi, S. Safi, S. Safiri, R. Sagar, A. Sahebkar, M. A. Sahraian, S. M. Sajadi, M. R. Salahshoor, P. Salamati, S. S. Zahabi, H. Salem, M. R. R. Salem, H. Salimzadeh, J. A. Salomon, I. Salz, Z. Samad, A. M. Samy, J. Sanabria, D. F. Santomauro, I. S. Santos, J. V. Santos, M. M. Santric-Milicevic, S. Y. I. Saraswathy, R. Sarmiento-Suárez, N. Sarrafzadegan, B. Sartorius, A. Sarveazad, B. Sathian, T. Sathish, D. Sattin, A. N. Sbarra, L. E. Schaeffer, S. Schiavolin, M. I. Schmidt, A. E. Schutte, D. C. Schwebel, F. Schwendicke, A. M. Senbeta, S. Senthilkumaran, S. G. Sepanlou, K. A. Shackelford, J. Shadid, S. Shahabi, A. A. Shaheen, M. A. Shaikh, A. S. Shalash, M. Shams-Beyranvand, M. Shamsizadeh, M. Shannawaz, K. Sharafi, F. Sharara, B. S. Sheena, A. Sheikhtaheri, R. S. Shetty, K. Shibuya, W. S. Shiferaw, M. Shigematsu, J. I. Shin, R. Shiri, R. Shirkoohi, M.

G. Shrime, K. Shuval, S. Siabani, I. D. Sigfusdottir, R. Sigurvinsdottir, J. P. Silva, K. E. Simpson, A. Singh, J. A. Singh, E. Skiadaresi, S. T. Skou, V. Y. Skryabin, E. Sobngwi, A. Sokhan, S. Soltani, R. J. D. Sorensen, J. B. Soriano, M. B. Sorrie, I. N. Soyiri, C. T. Sreeramareddy, J. D. Stanaway, B. A. Stark, S. C. Ştefan, C. Stein, C. Steiner, T. J. Steiner, M. A. Stokes, L. J. Stovner, J. L. Stubbs, A. Sudaryanto, M. B. Sufiyan, G. Sulo, I. Sultan, B. L. Sykes, D. O. Sylte, M. Szócska, R. Tabarés-Seisdedos, K. M. Tabb, S. K. Tadakamadla, A. Taherkhani, M. Tajdini, K. Takahashi, N. Taveira, W. L. Teagle, H. Teame, A. Tehrani-Banihashemi, B. F. Teklehaimanot, S. Terrason, Z. T. Tessema, K. R. Thankappan, A. M. Thomson, H. R. Tohidinik, M. Tonelli, R. Topor-Madry, A. E. Torre, M. Touvier, M. R. R. Tovani-Palone, B. X. Tran, R. Travillian, C. E. Troeger, T. C. Truelsen, A. C. Tsai, A. Tsatsakis, L. T. Car, S. Tyrovolas, R. Uddin, S. Ullah, E. A. Undurraga, B. Unnikrishnan, M. Vacante, A. Vakilian, P. R. Valdez, S. Varughese, T. J. Vasankari, Y. Vasseghian, N. Venketasubramanian, F. S. Violante, V. Vlassov, S. E. Vollset, A. Vongpradith, A. Vukovic, R. Vukovic, Y. Waheed, M. K. Walters, J. Wang, Y. Wang, Y.-P. Wang, J. L. Ward, A. Watson, J. Wei, R. G. Weintraub, D. J. Weiss, J. Weiss, R. Westerman, J. L. Whisnant, H. A. Whiteford, T. Wiangkham, K. E. Wiens, T. Wijeratne, L. B. Wilner, S. Wilson, B. Wojtyniak, C. D. A. Wolfe, E. E. Wool, A.-M. Wu, S. W. Hanson, H. Y. Wunrow, G. Xu, R. Xu, S. Yadgir, S. H. Y. Jabbari, K. Yamagishi, M. Yaminfirooz, Y. Yano, S. Yaya, V. Yazdi-Feyzabadi, J. A. Yearwood, T. Y. Yeheyis, Y. G. Yeshitila, P. Yip, N. Yonemoto, S.-J. Yoon, J. Y. Lebni, M. Z. Younis, T. P. Younker, Z. Yousefi, M. Yousefifard, T. Yousefinezhadi, A. Y. Yousuf, C. Yu, H. Yusefzadeh, T. Z. Moghadam, L. Zaki, S. B. Zaman, M. Zamani, M. Zamanian, H. Zandian, A. Zangeneh, M. S. Zastrozhin, K. A. Zewdie, Y. Zhang, Z.-J. Zhang, J. T. Zhao, Y. Zhao, P. Zheng, M. Zhou, A. Ziapour, S. R. M. Zimsen, M. Naghavi, C. J. L. Murray, Global burden of 369 diseases and injuries in 204 countries and territories, 1990–2019: a systematic analysis for the Global Burden of Disease Study 2019. *The Lancet* **396**, 1204–1222 (2020).

23. C. J. L. Murray, A. Y. Aravkin, P. Zheng, C. Abbafati, K. M. Abbas, M. Abbasi-Kangevari, F. Abd-Allah, A. Abdelalim, M. Abdollahi, I. Abdollahpour, K. H. Abegaz, H. Abolhassani, V. Aboyans, L. G. Abreu, M. R. M. Abrigo, A. Abualhasan, L. J. Abu-Raddad, A. I. Abushouk, M. Adabi, V. Adekanmbi, A. M. Adeoye, O. O. Adetokunboh, D. Adham, S. M. Advani, G. Agarwal, S. M. K. Aghamir, A. Agrawal, T. Ahmad, K. Ahmadi, M. Ahmadi, H. Ahmadieh, M. B. Ahmed, T. Y. Akalu, R. O. Akinyemi, T. Akinyemiju, B. Akombi, C. J. Akunna, F. Alahdab, Z. Al-Aly, K. Alam, S. Alam, T. Alam, F. M. Alanezi, T. M. Alanzi, B. wassihun Alemu, K. F. Alhabib, M. Ali, S. Ali, G. Alicandro, C. Alinia, V. Alipour, H. Alizade, S. M. Aljunid, F. Alla, P. Allebeck, A. Almasi-Hashiani, H. M. Al-Mekhlafi, J. Alonso, K. A. Altirkawi, M. Amini-Rarani, F. Amiri, D. A. Amugsi, R. Ancuceanu, D. Anderlini, J. A. Anderson, C. L. Andrei, T. Andrei, C. Angus, M. Anjomshoa, F. Ansari, A. Ansari-Moghaddam, I. C. Antonazzo, C. A. T. Antonio, C. M. Antony, E. Antriyandarti, D. Anvari, R. Anwer, S. C. Y. Appiah, J. Arabloo, M. Arab-Zozani, F. Ariani, B. Armoon, J. Ärnlöv, A. Arzani, M. Asadi-Aliabadi, A. A. Asadi-Pooya, C. Ashbaugh, M. Assmus, Z. Atafar, D. D. Atnafu, M. M. W. Atout, F. Ausloos, M. Ausloos, B. P. A. Quintanilla, G. Ayano, M. A. Ayanore, S. Azari, G. Azarian, Z. N. Azene, A. Badawi, A. D. Badiye, M. A. Bahrami, M. H. Bakhshaei, A. Bakhtiari, S. M. Bakkannavar, A. Baldasseroni, K. Ball, S. H. Ballew, D. Balzi, M. Banach, S. K. Banerjee, A. B. Bante, A. G. Baraki, S. L. Barker-Collo, T. W. Bärnighausen, L. H. Barrero, C. M.

Barthelemy, L. Barua, S. Basu, B. T. Baune, M. Bayati, J. S. Becker, N. Bedi, E. Beghi, Y. Béjot, M. L. Bell, F. B. Bennitt, I. M. Bensenor, K. Berhe, A. E. Berman, A. S. Bhagavathula, R. Bhageerathy, N. Bhala, D. Bhandari, K. Bhattacharyya, Z. A. Bhutta, A. Bijani, B. Bikbov, M. S. B. Sayeed, A. Biondi, B. M. Birihane, C. Bisignano, R. K. Biswas, H. Bitew, S. Bohlouli, M. Bohluli, A. S. Boon-Dooley, G. Borges, A. M. Borzì, S. Borzouei, C. Bosetti, S. Boufous, D. Braithwaite, N. J. K. Breitborde, S. Breitner, H. Brenner, P. S. Briant, A. N. Briko, N. I. Briko, G. B. Britton, D. Bryazka, B. R. Bumgarner, K. Burkart, R. T. Burnett, S. B. Nagaraja, Z. A. Butt, F. L. C. dos Santos, L. E. Cahill, L. L. A. Cámera, I. R. Campos-Nonato, R. Cárdenas, G. Carreras, J. J. Carrero, F. Carvalho, J. M. Castaldelli-Maia, C. A. Castañeda-Orjuela, G. Castelpietra, F. Castro, K. Causey, C. R. Cederroth, K. M. Cercy, E. Cerin, J. S. Chandan, K.-L. Chang, F. J. Charlson, V. K. Chattu, S. Chaturvedi, N. Cherbuin, O. Chimed-Ochir, D. Y. Cho, J.-Y. J. Choi, H. Christensen, D.-T. Chu, M. T. Chung, S.-C. Chung, F. M. Cicuttini, L. G. Ciobanu, M. Cirillo, T. K. D. Classen, A. J. Cohen, K. Compton, O. R. Cooper, V. M. Costa, E. Cousin, R. G. Cowden, D. H. Cross, J. A. Cruz, S. M. A. Dahlawi, A. A. M. Damasceno, G. Damiani, L. Dandona, R. Dandona, W. J. Dangel, A.-K. Danielsson, P. I. Dargan, A. M. Darwesh, A. Daryani, J. K. Das, R. D. Gupta, J. das Neves, C. A. Dávila-Cervantes, D. V. Davitoiu, D. D. Leo, L. Degenhardt, M. DeLang, R. P. Dellavalle, F. M. Demeke, G. T. Demoz, D. G. Demsie, E. Denova-Gutiérrez, N. Dervenis, G. P. Dhungana, M. Dianatinasab, D. D. da Silva, D. Diaz, Z. S. D. Forooshani, S. Djalalinia, H. T. Do, K. Dokova, F. Dorostkar, L. Doshmangir, T. R. Driscoll, B. B. Duncan, A. R. Duraes, A. W. Eagan, D. Edvardsson, N. E. Nahas, I. E. Sayed, M. E. Tantawi, I. Elbarazi, I. Y. Elgendy, S. I. El-Jaafary, I. R. Elyazar, S. Emmons-Bell, H. E. Erskine, S. Eskandarieh, S. Esmaeilnejad, A. Esteghamati, K. Estep, A. Etemadi, A. E. Etisso, J. Fanzo, M. Farahmand, M. Fareed, R. Faridnia, A. Farioli, A. Faro, M. Faruque, F. Farzadfar, N. Fattahi, M. Fazlzadeh, V. L. Feigin, R. Feldman, S.-M. Fereshtehnejad, E. Fernandes, G. Ferrara, A. J. Ferrari, M. L. Ferreira, I. Filip, F. Fischer, J. L. Fisher, L. S. Flor, N. A. Foigt, M. O. Folayan, A. A. Fomenkov, L. M. Force, M. Foroutan, R. C. Franklin, M. Freitas, W. Fu, T. Fukumoto, J. M. Furtado, M. M. Gad, E. Gakidou, S. Gallus, A. L. Garcia-Basteiro, W. M. Gardner, B. S. Geberemariyam, A. A. A. A. Gebreslassie, A. Geremew, A. G. Hayoon, P. W. Gething, M. Ghadimi, K. Ghadiri, F. Ghaffarifar, M. Ghafourifard, F. Ghamari, A. Ghashghaee, H. Ghiasvand, N. Ghith, A. Gholamian, R. Ghosh, P. S. Gill, T. G. G. Ginindza, G. Giussani, E. V. Gnedovskaya, S. Goharinezhad, S. V. Gopalani, G. Gorini, H. Goudarzi, A. C. Goulart, F. Greaves, M. Grivna, G. Grosso, M. I. M. Gubari, H. C. Gugnani, R. A. Guimarães, R. A. Guled, G. Guo, Y. Guo, R. Gupta, T. Gupta, B. Haddock, N. Hafezi-Nejad, A. Hafiz, A. Haj-Mirzaian, A. Haj-Mirzaian, B. J. Hall, I. Halvaei, R. R. Hamadeh, S. Hamidi, M. S. Hammer, G. J. Hankey, H. Haririan, J. M. Haro, A. I. Hasaballah, M. M. Hasan, E. Hasanpoor, A. Hashi, S. Hassanipour, H. Hassankhani, R. J. Havmoeller, S. I. Hay, K. Hayat, G. Heidari, R. Heidari-Soureshjani, H. J. Henrikson, M. E. Herbert, C. Herteliu, F. Heydarpour, T. R. Hird, H. W. Hoek, R. Holla, P. Hoogar, H. D. Hosgood, N. Hossain, M. Hosseini, M. Hosseinzadeh, M. Hostiuc, S. Hostiuc, M. Househ, M. Hsairi, V. C. Hsieh, G. Hu, K. Hu, T. M. Huda, A. Humayun, C. K. Huynh, B.-F. Hwang, V. C. Iannucci, S. E. Ibitoye, N. Ikeda, K. S. Ikuta, O. S. Ilesanmi, I. M. Ilic, M. D. Ilic, L. R. Inbaraj, H. Ippolito, U. Iqbal, S. S. N. Irvani, C. M. S. Irvine, M. M. Islam, S. M. S. Islam, H. Iso, R. Q. Ivers, C. C. D. Iwu, C. J. Iwu, I. O. Iyamu, J. Jaafari, K. H. Jacobsen, H. Jafari, M. Jafarinia, M. A. Jahani, M. Jakovljevic, F. Jalilian, S. L. James, H. Janjani, T. Javaheri, J. Javidnia, P. Jeemon, E.

Jenabi, R. P. Jha, V. Jha, J. S. Ji, L. Johansson, O. John, Y. O. John-Akinola, C. O. Johnson, J. B. Jonas, F. Joukar, J. J. Jozwiak, M. Jürisson, A. Kabir, Z. Kabir, H. Kalani, R. Kalani, L. R. Kalankesh, R. Kalhor, T. Kanchan, N. Kapoor, B. K. Matin, A. Karch, M. A. Karim, G. M. Kassa, S. V. Katikireddi, G. A. Kayode, A. K. Karyani, P. N. Keiyoro, C. Keller, L. Kemmer, P. J. Kendrick, N. Khalid, M. Khammarnia, E. A. Khan, M. Khan, K. Khatab, M. M. Khater, M. N. Khatib, M. Khayamzadeh, S. Khazaei, C. Kieling, Y. J. Kim, R. W. Kimokoti, A. Kisa, S. Kisa, M. Kivimäki, L. D. Knibbs, A. K. S. Knudsen, J. M. Kocarnik, S. Kochhar, J. A. Kopec, V. A. Korshunov, P. A. Koul, A. Koyanagi, M. U. G. Kraemer, K. Krishan, K. J. Krohn, H. Kromhout, B. K. Defo, G. A. Kumar, V. Kumar, O. P. Kurmi, D. Kusuma, C. L. Vecchia, B. Lacey, D. K. Lal, R. Lalloo, T. Lallukka, F. H. Lami, I. Landires, J. J. Lang, S. M. Langan, A. O. Larsson, S. Lasrado, P. Lauriola, J. V. Lazarus, P. H. Lee, S. W. H. Lee, K. E. LeGrand, J. Leigh, M. Leonardi, H. Lescinsky, J. Leung, M. Levi, S. Li, L.-L. Lim, S. Linn, S. Liu, S. Liu, Y. Liu, J. Lo, A. D. Lopez, J. C. F. Lopez, P. D. Lopukhov, S. Lorkowski, P. A. Lotufo, A. Lu, A. Lugo, E. R. Maddison, P. W. Mahasha, M. M. Mahdavi, M. Mahmoudi, A. Majeed, A. Maleki, S. Maleki, R. Malekzadeh, D. C. Malta, A. A. Mamun, A. L. Manda, H. Manguerra, F. Mansour-Ghanaei, B. Mansouri, M. A. Mansournia, A. M. M. Herrera, J. C. Maravilla, A. Marks, R. V. Martin, S. Martini, F. R. Martins-Melo, A. Masaka, S. Z. Masoumi, M. R. Mathur, K. Matsushita, P. K. Maulik, C. McAlinden, J. J. McGrath, M. McKee, M. M. Mehndiratta, F. Mehri, K. M. Mehta, Z. A. Memish, W. Mendoza, R. G. Menezes, E. W. Mengesha, A. Mereke, S. T. Mereta, A. Meretoja, T. J. Meretoja, T. Mestrovic, B. Miazgowski, T. Miazgowski, I. M. Michalek, T. R. Miller, E. J. Mills, G. K. Mini, M. Miri, A. Mirica, E. M. Mirrakhimov, H. Mirzaei, M. Mirzaei, R. Mirzaei, M. Mirzaei-Alavijeh, A. T. Misganaw, P. Mithra, B. Moazen, D. K. Mohammad, Y. Mohammad, N. M. G. Mezerji, A. Mohammadian-Hafshejani, N. Mohammadifard, R. Mohammadpourhodki, A. S. Mohammed, H. Mohammed, J. A. Mohammed, S. Mohammed, A. H. Mokdad, M. Molokhia, L. Monasta, M. D. Mooney, G. Moradi, M. Moradi, M. Moradi-Lakeh, R. Moradzadeh, P. Moraga, L. Morawska, J. Morgado-da-Costa, S. D. Morrison, A. Mosapour, J. F. Mosser, S. Mouodi, S. M. Mousavi, A. M. Khaneghah, U. O. Mueller, S. Mukhopadhyay, E. C. Mullany, K. I. Musa, S. Muthupandian, A. F. Nabhan, M. Naderi, A. J. Nagarajan, G. Nagel, M. Naghavi, B. Naghshtabrizi, M. D. Naimzada, F. Najafi, V. Nangia, J. R. Nansseu, M. Naserbakht, V. C. Nayak, I. Negoi, J. W. Ngunjiri, C. T. Nguyen, H. L. T. Nguyen, M. Nguyen, Y. T. Nigatu, R. Nikbakhsh, M. R. Nixon, C. A. Nnaji, S. Nomura, B. Norrving, J. J. Noubiap, C. Nowak, V. Nunez-Samudio, A. Oţoiu, B. Oancea, C. M. Odell, F. A. Ogbo, I.-H. Oh, E. W. Okunga, M. Oladnabi, A. T. Olagunju, B. O. Olusanya, J. O. Olusanya, M. O. Omer, K. L. Ong, O. E. Onwujekwe, H. M. Orpana, A. Ortiz, O. Osarenotor, F. B. Osei, S. M. Ostroff, N. Otstavnov, S. S. Otstavnov, S. Øverland, M. O. Owolabi, M. P. A, J. R. Padubidri, R. Palladino, S. Panda-Jonas, A. Pandey, C. D. H. Parry, M. Pasovic, D. K. Pasupula, S. K. Patel, M. Pathak, S. B. Patten, G. C. Patton, H. P. Toroudi, A. E. Peden, A. Pennini, V. C. F. Pepito, E. K. Peprah, D. M. Pereira, K. Pesudovs, H. Q. Pham, M. R. Phillips, C. Piccinelli, T. M. Pilz, M. A. Piradov, M. Pirsaheb, D. Plass, S. Polinder, K. R. Polkinghorne, C. D. Pond, M. J. Postma, H. Pourjafar, F. Pourmalek, A. Poznańska, S. I. Prada, V. Prakash, D. R. A. Pribadi, E. Pupillo, Z. Q. Syed, M. Rabiee, N. Rabiee, A. Radfar, A. Rafiee, A. Raggi, M. A. Rahman, A. Rajabpour-Sanati, F. Rajati, I. Rakovac, P. Ram, K. Ramezanzadeh, C. L. Ranabhat, P. C. Rao, S. J. Rao, V. Rashedi, P. Rathi, D. L. Rawaf, S. Rawaf, L. Rawal, R. Rawassizadeh, R. Rawat,

C. Razo, S. B. Redford, R. C. Reiner, M. B. Reitsma, G. Remuzzi, V. Renjith, A. M. N. Renzaho, S. Resnikoff, N. Rezaei, N. Rezaei, A. Rezapour, P.-A. Rhinehart, S. M. Riahi, D. C. Ribeiro, D. Ribeiro, J. Rickard, J. A. Rivera, N. L. S. Roberts, S. Rodríguez-Ramírez, L. Roever, L. Ronfani, R. Room, G. Roshandel, G. A. Roth, D. Rothenbacher, E. Rubagotti, G. M. Rwegerera, S. Sabour, P. S. Sachdev, B. Saddik, E. Sadeghi, M. Sadeghi, R. Saeedi, S. S. Moghaddam, Y. Safari, S. Safi, S. Safiri, R. Sagar, A. Sahebkar, S. M. Sajadi, N. Salam, P. Salamati, H. Salem, M. R. R. Salem, H. Salimzadeh, O. M. Salman, J. A. Salomon, Z. Samad, H. S. Kafil, E. Z. Sambala, A. M. Samy, J. Sanabria, T. G. Sánchez-Pimienta, D. F. Santomauro, I. S. Santos, J. V. Santos, M. M. Santric-Milicevic, S. Y. I. Saraswathy, R. Sarmiento-Suárez, N. Sarrafzadegan, B. Sartorius, A. Sarveazad, B. Sathian, T. Sathish, D. Sattin, S. Saxena, L. E. Schaeffer, S. Schiavolin, M. P. Schlaich, M. I. Schmidt, A. E. Schutte, D. C. Schwebel, F. Schwendicke, A. M. Senbeta, S. Senthilkumaran, S. G. Sepanlou, B. Serdar, M. L. Serre, J. Shadid, O. Shafaat, S. Shahabi, A. A. Shaheen, M. A. Shaikh, A. S. Shalash, M. Shams-Beyranvand, M. Shamsizadeh, K. Sharafi, A. Sheikh, A. Sheikhtaheri, K. Shibuya, K. D. Shield, M. Shigematsu, J. I. Shin, M.-J. Shin, R. Shiri, R. Shirkoohi, K. Shuval, S. Siabani, R. Sierpinski, I. D. Sigfusdottir, R. Sigurvinsdottir, J. P. Silva, K. E. Simpson, J. A. Singh, P. Singh, E. Skiadaresi, S. T. Skou, V. Y. Skryabin, E. U. R. Smith, A. Soheili, S. Soltani, M. Soofi, R. J. D. Sorensen, J. B. Soriano, M. B. Sorrie, S. Soshnikov, I. N. Soyiri, C. N. Spencer, A. Spotin, C. T. Sreeramareddy, V. Srinivasan, J. D. Stanaway, C. Stein, D. J. Stein, C. Steiner, L. Stockfelt, M. A. Stokes, K. Straif, J. L. Stubbs, M. B. Sufiyan, H. A. R. Suleria, R. S. Abdulkader, G. Sulo, I. Sultan, Ł. Szumowski, R. Tabarés-Seisdedos, K. M. Tabb, T. Tabuchi, A. Taherkhani, M. Tajdini, K. Takahashi, J. S. Takala, A. T. Tamiru, N. Taveira, A. Tehrani-Banihashemi, M.-H. Temsah, G. A. Tesema, Z. T. Tessema, G. D. Thurston, M. V. Titova, H. R. Tohidinik, M. Tonelli, R. Topor-Madry, F. Topouzis, A. E. Torre, M. Touvier, M. R. R. Tovani-Palone, B. X. Tran, R. Travillian, A. Tsatsakis, L. T. Car, S. Tyrovolas, R. Uddin, C. D. Umeokonkwo, B. Unnikrishnan, E. Upadhyay, M. Vacante, P. R. Valdez, A. van Donkelaar, T. J. Vasankari, Y. Vasseghian, Y. Veisani, N. Venketasubramanian, F. S. Violante, V. Vlassov, S. E. Vollset, T. Vos, R. Vukovic, Y. Waheed, M. T. Wallin, Y. Wang, Y.-P. Wang, A. Watson, J. Wei, M. Y. W. Wei, R. G. Weintraub, J. Weiss, A. Werdecker, J. J. West, R. Westerman, J. L. Whisnant, H. A. Whiteford, K. E. Wiens, C. D. A. Wolfe, S. S. Wozniak, A.-M. Wu, J. Wu, S. W. Hanson, G. Xu, R. Xu, S. Yadgir, S. H. Y. Jabbari, K. Yamagishi, M. Yaminfirooz, Y. Yano, S. Yaya, V. Yazdi-Feyzabadi, T. Y. Yeheyis, C. S. Yilgwan, M. T. Yilma, P. Yip, N. Yonemoto, M. Z. Younis, T. P. Younker, B. Yousefi, Z. Yousefi, T. Yousefinezhadi, A. Y. Yousuf, C. Yu, H. Yusefzadeh, T. Z. Moghadam, M. Zamani, M. Zamanian, H. Zandian, M. S. Zastrozhin, Y. Zhang, Z.-J. Zhang, J. T. Zhao, X.-J. G. Zhao, Y. Zhao, M. Zhou, A. Ziapour, S. R. M. Zimsen, M. Brauer, A. Afshin, S. S. Lim, Global burden of 87 risk factors in 204 countries and territories, 1990–2019: a systematic analysis for the Global Burden of Disease Study 2019. *The Lancet* **396**, 1223–1249 (2020).
